# The impact of SARS-CoV-2 vaccines on antibody responses in the general population in the United Kingdom

**DOI:** 10.1101/2021.04.22.21255911

**Authors:** Jia Wei, Nicole Stoesser, Philippa C. Matthews, Ruth Studley, Iain Bell, John I Bell, John N Newton, Jeremy Farrar, Ian Diamond, Emma Rourke, Alison Howarth, Brian D. Marsden, Sarah Hoosdally, E Yvonne Jones, David I Stuart, Derrick W .Crook, Tim E. A. Peto, Koen B. Pouwels, David W. Eyre, A. Sarah Walker, COVID-19 Infection Survey team

## Abstract

Real-world data on antibody response post-vaccination in the general population are limited. 45,965 adults in the UK’s national COVID-19 Infection Survey receiving Pfizer-BioNTech or Oxford-AstraZeneca vaccines had 111,360 anti-spike IgG measurements. Without prior infection, seroconversion rates and quantitative antibody levels post single dose were lower in older individuals, especially >60y. Two doses achieved high responses across all ages, particularly increasing seroconversion in older people, to similar levels to those achieved after prior infection followed by a single dose. Antibody levels rose more slowly and to lower levels with Oxford-AstraZeneca vs Pfizer-BioNTech, but waned following a single Pfizer-BioNTech dose. Latent class models identified four responder phenotypes: older people, males, and those having long-term health conditions were more commonly ‘low responders’. Where supplies are limited, vaccines should be prioritised for those not previously infected, and second doses to individuals >60y. Further data on the relationship between vaccine-mediated protection and antibody responses are needed.

## Introduction

Multiple vaccines have been developed which offer protection against COVID-19 by generating immune responses against the spike antigen of SARS-CoV-2. On 8 December 2020, the United Kingdom (UK) started its national vaccination programme, shortly after becoming the first country to approve the Pfizer-BioNTech BNT162b2 vaccine^1^, followed by the approval of the Oxford-AstraZeneca ChAdOx1 nCoV-19 vaccine, first used outside a clinical trial on 4 January 2021^2^. Both vaccines have been widely used in the UK.

These vaccines were initially administered to priority groups identified by the Joint Committee on Vaccination and Immunisation, including elderly people in care homes, people over 80 years old, healthcare workers, and clinically vulnerable people, and then offered to the rest of the adult population in decreasing age order^3^. To maximise initial coverage, in early January the dosing interval was extended to 12 weeks for all vaccines, regardless of licensed dosing schedule. To 6 April 2021, over 31.7 million people (60.2% of the total population aged 18 and over) have been given a first dose, and 5.7 million people (10.8% of the total population aged 18 and over) have received two doses of vaccine (https://coronavirus.data.gov.uk/details/vaccinations).

The efficacy of the Oxford-AstraZeneca and Pfizer-BioNTech vaccines against symptomatic laboratory-confirmed COVID-19 infection has been reported in large randomized controlled clinical trials as 52% (95%CI 30-86%) after the first dose and 95% (90-98%) after the second dose of the Pfizer-BioNTech vaccine^4^, and 70% (55-81%) after the second dose of the Oxford-AstraZeneca vaccine^5^. Several studies have examined the immunogenicity of vaccines in healthcare workers, typically the earliest groups to be vaccinated. A study of 3610 healthcare workers found that 99.5% and 97.1% seroconverted after a single dose of the Pfizer-BioNTech and Oxford-AstraZeneca vaccines respectively, and that higher quantitative IgG levels were achieved in previously infected seropositive individuals^6^. Other studies have also found that, compared to seronegative individuals, a single dose of Pfizer-BioNTech elicited higher antibody levels in previously seropositive individuals, comparable to the levels achieved after two doses of vaccines in seronegative individuals^7–9^. Outside of trials, there is limited data on post-vaccine antibody responses in other groups, especially older adults who were also underrepresented in the Oxford-AstraZeneca trial^5^. One small study of 185 individuals aged >70 years showed very high seropositivity after one or two Pfizer-BioNTech doses^10^. A second study of 100 individuals aged 80-100 years showed almost universal high antibody responses 3 weeks after a single dose of Pfizer-BioNtech, with spike-specific cellular responses in 63%^11^. However, the representativeness of these small cohorts is unclear.

Real-world data can provide information on populations who may not participate in clinical trials as well as assessing the efficacy of interventions as actually deployed. Therefore, we used the UK’s national COVID-19 Infection Survey (CIS) (ISRCTN21086382), which includes a representative sample of households and has longitudinal follow-up, to study population-wide vaccine immunogenicity. We investigated anti-trimeric spike IgG antibody responses after vaccination by time since vaccination, considering vaccine type (Pfizer-BioNTech or Oxford-AstraZeneca), the number of doses received, and whether there was evidence of prior SARS-CoV-2 infection. We also assessed how demographic factors, including age, sex, and ethnicity, impact post-vaccine antibody responses.

## Results

45,965 participants aged ≥16 years from the general population who were first vaccinated between 8 December 2020 and 6 April 2021 contributed a total of 111,360 SARS-CoV-2 anti-spike IgG measurements taken at any point between 91 days before first vaccination date through to 6 April 2021. The median (IQR) age was 64 (54-71) years and 25,330 (55.1%) were female. 2,745 (6.0%) were healthcare workers, and 15,334 (33.4%) had a long-term health condition (**Table S1**). 5,834 (12.7%) participants with a SARS-CoV-2 PCR-positive study nose/throat swab or anti-spike IgG positive study antibody result at any time prior to vaccination were considered to have been previously infected with SARS-CoV-2, irrespective of whether they had reported previous symptoms or not. Using this definition, 3,767 (8.2%) and 2,067 (4.5%) previously infected participants received one dose of Oxford-AstraZeneca or Pfizer-BioNTech, respectively. 23,368 (50.8%), 14,894 (32.4%), and 1,869 (4.1%) participants without evidence of prior infection received one dose of Oxford-AstraZeneca, one dose of Pfizer-BioNTech, and two doses of Pfizer-BioNTech, respectively. Among 1,869 (4.1%) participants without evidence of prior infection who received two doses of Pfizer-BioNTech, the median (IQR) duration between two doses was 31 (21-47) days, with 1,020 (54.6%) ≤31 days (**Figure 1, Table S1**). Participant characteristics varied across the different vaccination groups, generally reflecting the prioritisation, with proportionately more healthcare workers and the oldest individuals having received two doses of Pfizer-BioNTech.

**Figure 1.**
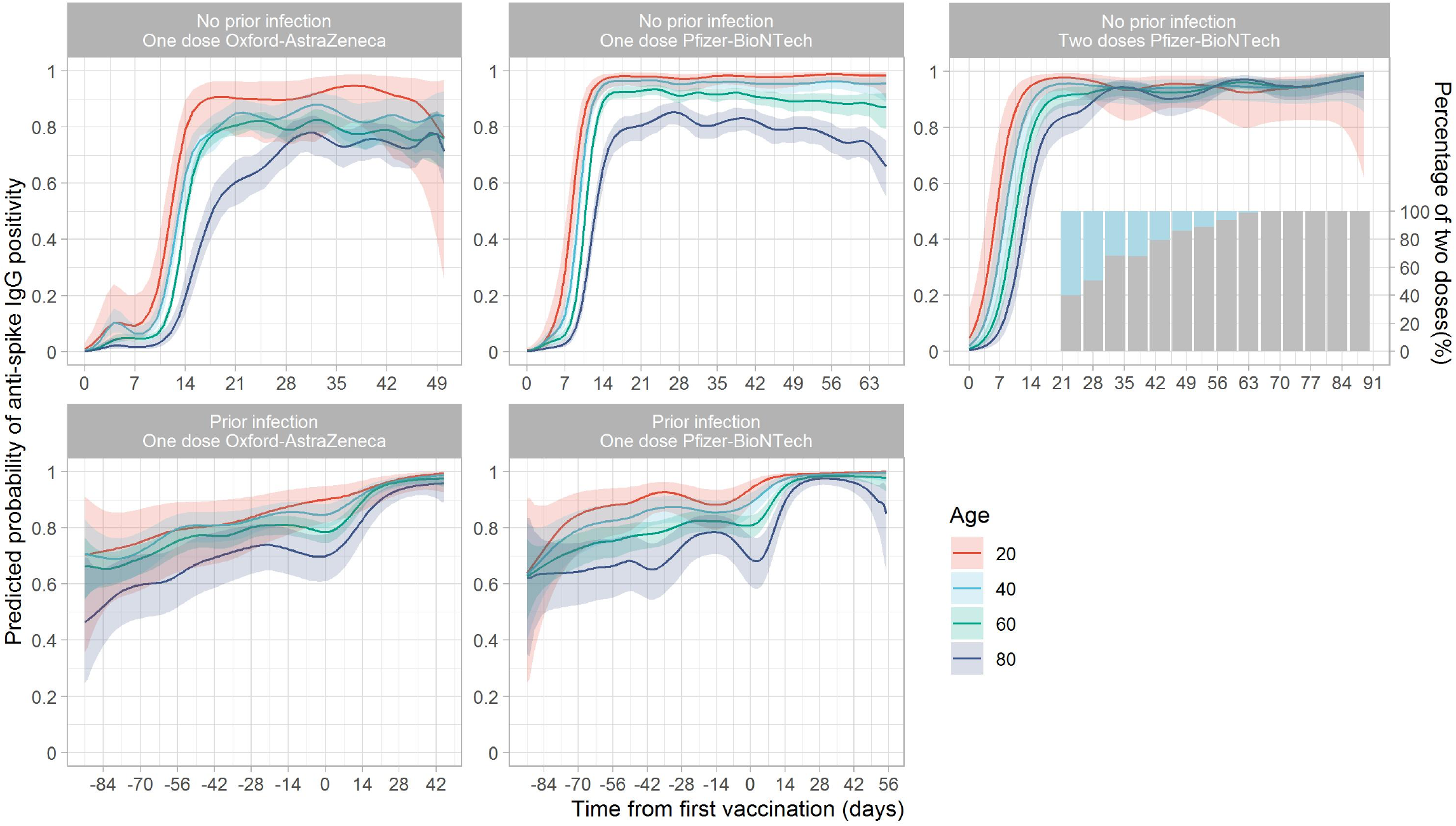
Predicted probability of anti-spike IgG positivity by time from first vaccination, according to vaccine type and prior infection status. Line colour indicates antibody response predicted for ages 20, 40, 60, and 80 years (see **Figure S1** for full model across all ages and **Figure S2** for comparisons of vaccine type by age). The bars in ‘Two doses Pfizer-BioNTech’ plot show the percentage of people having had two doses of vaccines by each timepoint (Gray: had two doses; Blue: had only one dose).Different x-axis scales reflect different durations of follow-up post-vaccination in the different cohorts.

### Antibody positivity after vaccination

In models of positive vs. negative post-vaccine antibody responses in participants without evidence of prior infection, the estimated percentage with a positive anti-spike IgG result after vaccination increased over time and varied significantly by age (**Figures 1, S1, S2**; observed numbers/percentages in **Figures S3-S7**). Older participants had lower seropositivity rates than younger participants after receiving a single dose of Oxford-AstraZeneca or Pfizer-BioNTech, with most marked differences with increasing age over 60 years. For example, the estimated percentage of seropositive 80-year-olds was 74% (95%CI 66-80%) and 85% (80-89%) 28 days after first vaccination with Oxford-AstraZeneca or Pfizer-BioNTech vaccine, respectively, compared with 79% (75-83%) and 91% (89-93%) for 60-year-olds and 84% (76-89%) and 95% (92-97%) for 40-year-olds (**Table S2**). In contrast, two doses of Pfizer-BioNTech vaccine achieved >90% seropositivity 28-72 days after the first vaccination regardless of age, although there was some evidence of waning in those only receiving a first Pfizer-BioNTech doses at older ages. There was no evidence of differences in seropositivity rates 14-42 days post-first vaccine in those of younger ages (e.g. 20, 40 years) receiving one dose or two doses of Pfizer-BioNTech but greater rates of seroconversion were seen in older individuals (e.g. 80 years) receiving two doses (**Figure S2**). There was no evidence of declines following first vaccine dose in older individuals receiving a single dose of Oxford-AstraZeneca.

In participants with prior evidence of infection, before vaccination the predicted probability of being seropositive showed an inverse association with age, e.g. on the day of vaccination it was 90% (95%CI 82-95%) for 20-year-olds, 85% (80-88%) for 40-year-olds, 78% (75-82%) for 60-year-olds, and 70% (61-78%) for 80-year-olds receiving Oxford-AstraZeneca (**Table S2**; same trend for Pfizer-BioNTech). A high percentage of participants achieved positive antibody responses 28 days after vaccination (≥94%) regardless of age and vaccine given, similar to the positivity rate in participants without prior infection who received two doses of Pfizer-BioNTech (**Figure S2**).

### Associations with initial antibody response in those without evidence of prior infection

28,144 participants had an anti-spike IgG measurement 14-60 days after their first vaccination, of whom 24,977 (88.7%) had no evidence of prior infection and were included in analyses of associations with antibody positivity. 20,505 (82.1%) had a positive anti-spike IgG result. Age, sex, vaccine type, ethnicity, social deprivation, healthcare roles, and long-term health conditions were associated with seropositivity after vaccination (**Table 1**). Consistent with **Figure 1**, initial anti-spike IgG positivity decreased with older age, and the association was not linear, with the positivity rate dropping faster after 75 years (**Figure 2A&B**). There was evidence of effect modification between age and sex, such that at younger ages (30-55y) similar rates of seroconversion were seen in males and females, but at older ages (>60y) men were less likely to seroconvert (**Figure 2A**, interaction p=0.01). For example, in 40 year-olds the adjusted OR (aOR) for seroconversion in males vs. females was 0.91 (95%CI 0.68-1.22), but was 0.65 (95%CI 0.59-0.72) for 70-year-olds. Seroconversion by 60 days was less common following Oxford-AstraZeneca than after Pfizer-BioNTech (aOR=0.47, 95%CI 0.44-0.51, p<0.001), while receiving two doses of Pfizer-BioNTech increased seroconversion compared to one Pfizer-BioNTech dose (aOR=2.11, 95%CI 1.69-2.66, p<0.001). Patient-facing healthcare workers were more likely to be anti-spike IgG positive by 60 days post-vaccination (aOR=1.63, 95%CI 1.29-2.08, p<0.001) and participants who had long-term health conditions were less likely (aOR=0.64, 95%CI 0.60-0.69, p<0.001). There was evidence of greater seropositivity post-vaccination in participants from non-white ethnic groups (aOR=1.54 95%CI 1.27-1.90, p<0.001). A 10-unit increase in deprivation percentile (i.e. decrease in deprivation) resulted in higher seropositivity post-vaccination (aOR=1.28, 95%CI 1.13-1.46, p<0.001). There was no evidence of independent associations between antibody positivity and household size or working in social care or long-term care facilities.

**Table 1.**
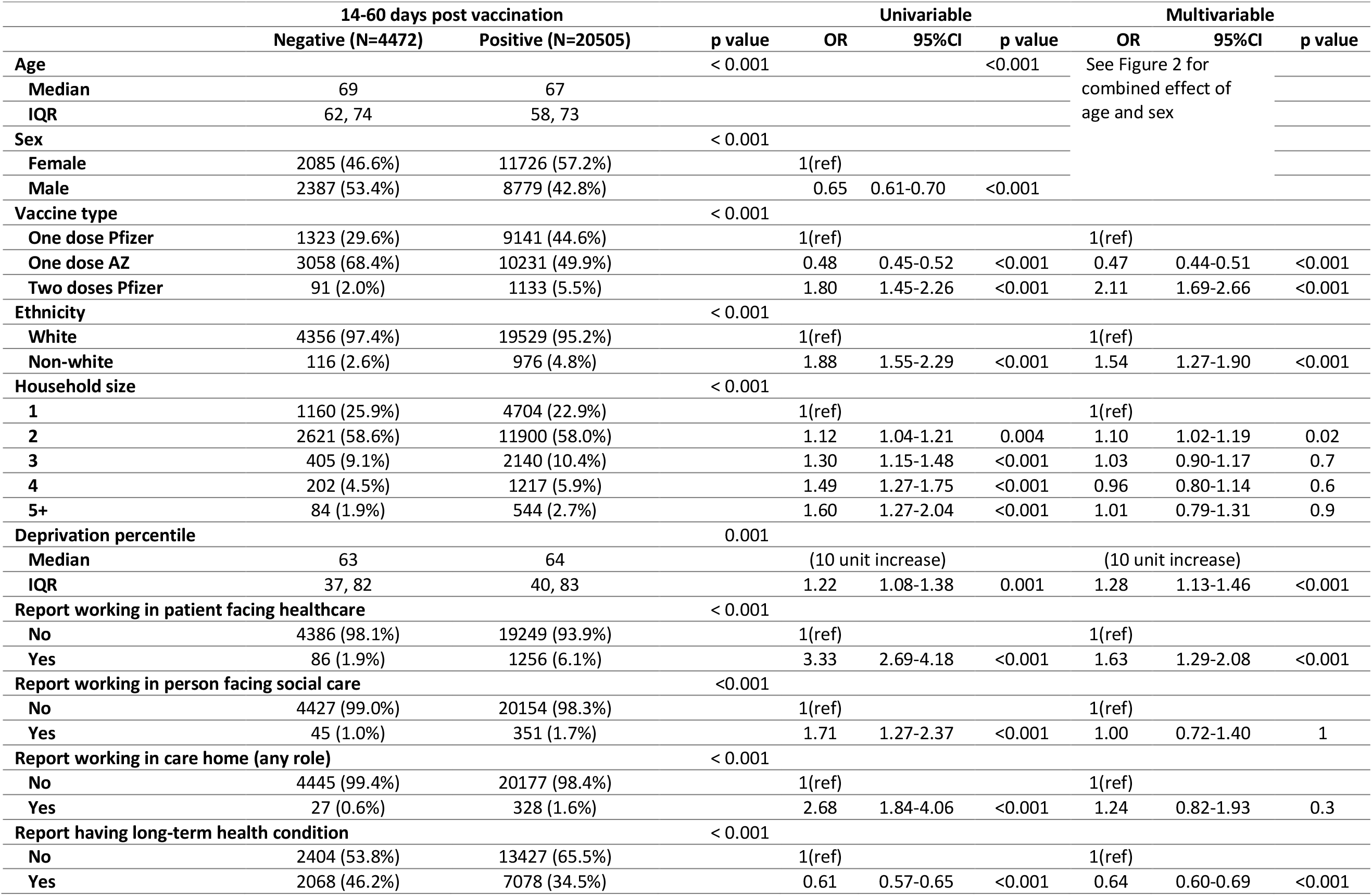
Predictors of antibody positivity 14-60 days post first vaccination in participants without evidence of prior infection from univariable and multivariable logistic regression models. AZ: Oxford-AstraZeneca vaccine. Pfizer: Pfizer-BioNTech vaccine.

**Figure 2.**
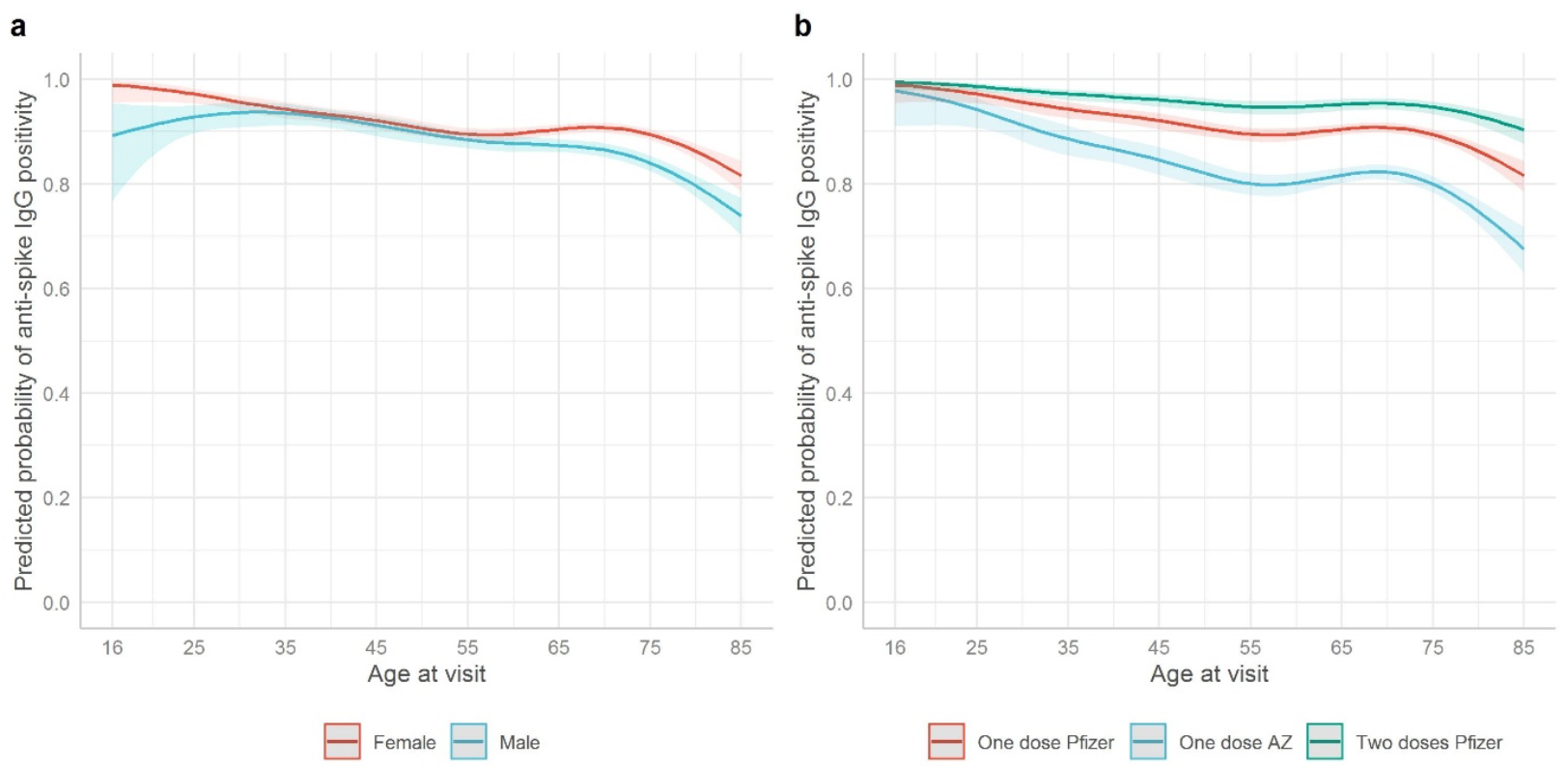
Predicted probability of spike IgG positivity 14-60 days after first vaccination in participants without evidence of prior infection from multivariable model. Panel A: Predicted probability of anti-spike IgG positivity by age and sex interaction. Panel B: Predicted probability of anti-spike IgG positivity by age and vaccine type. Age was fitted using natural cubic spline with four internal knots placed at 20^th^, 40^th^, 60^th^, and 80^th^ percentile (30, 44, 57, 71 years) and two boundary knots at 5^th^ and 95^th^ percentile (19, 82 years). Test for interaction between sex and age p=0.02.Plotted at reference category for other variables (Pfizer one dose (Panel A)/female (Panel B), white ethnicity, IMD=55, household size=1, did not work in patient-facing healthcare or social care, did not work in a care home, no long-term health condition).

### Quantitative antibody response after vaccination

In participants without evidence of prior infection, changes in quantitative anti-spike IgG levels followed similar patterns to binary IgG positivity post-vaccination (**Figures 3, 4, S8**). After receiving a single dose of Pfizer-BioNTech or Oxford-AstraZeneca vaccine, older participants reached lower peak levels and levels rose more slowly than in those of younger ages. Participants who received a single dose of Pfizer-BioNTech vaccine initially achieved higher anti-spike IgG levels than those who received Oxford-AstraZeneca vaccine. For example, 28 days after receiving Oxford-AstraZeneca vaccine and Pfizer-BioNTech vaccine, the IgG levels were 73 (95%CI 65-81) and 113 (104-123) for 80-year-olds, 94 (87-100) and 163 (153-175) for 60-year-olds, 113 (99-129) and 236 (214-261) for 40-year-olds, and 127 (94-171) and 334 (266-420) ng/ml equivalents for 20-year-olds, respectively (**Table S2**). The rate of increase in antibody levels was also slightly slower following the Oxford-AstraZeneca vaccine, e.g. the estimated mean time to reaching the threshold for antibody positivity post-first vaccine in 40-year-olds was around 10 days after receiving Pfizer-BioNTech but 14 days after receiving Oxford-AstraZeneca (**Figure 3**). However, antibody levels gradually decreased from ∼35 days post-vaccination in participants receiving a single dose of Pfizer-BioNTech (**Figure 3**), while there was no evidence of decrease in those receiving a single Oxford-AstraZeneca dose up to 49 days post-vaccination. Overall, differences in mean antibody levels between single doses of the two vaccines attenuated over time, particularly at older ages; for example, at 49 days, an 80-year-old receiving a single Pfizer-BioNTech dose had similar antibody levels (81, 95%CI 74-89) to one receiving a single Oxford-AstraZeneca dose (73, 95%CI 62-87) (**Table S2**).

**Figure 3.**
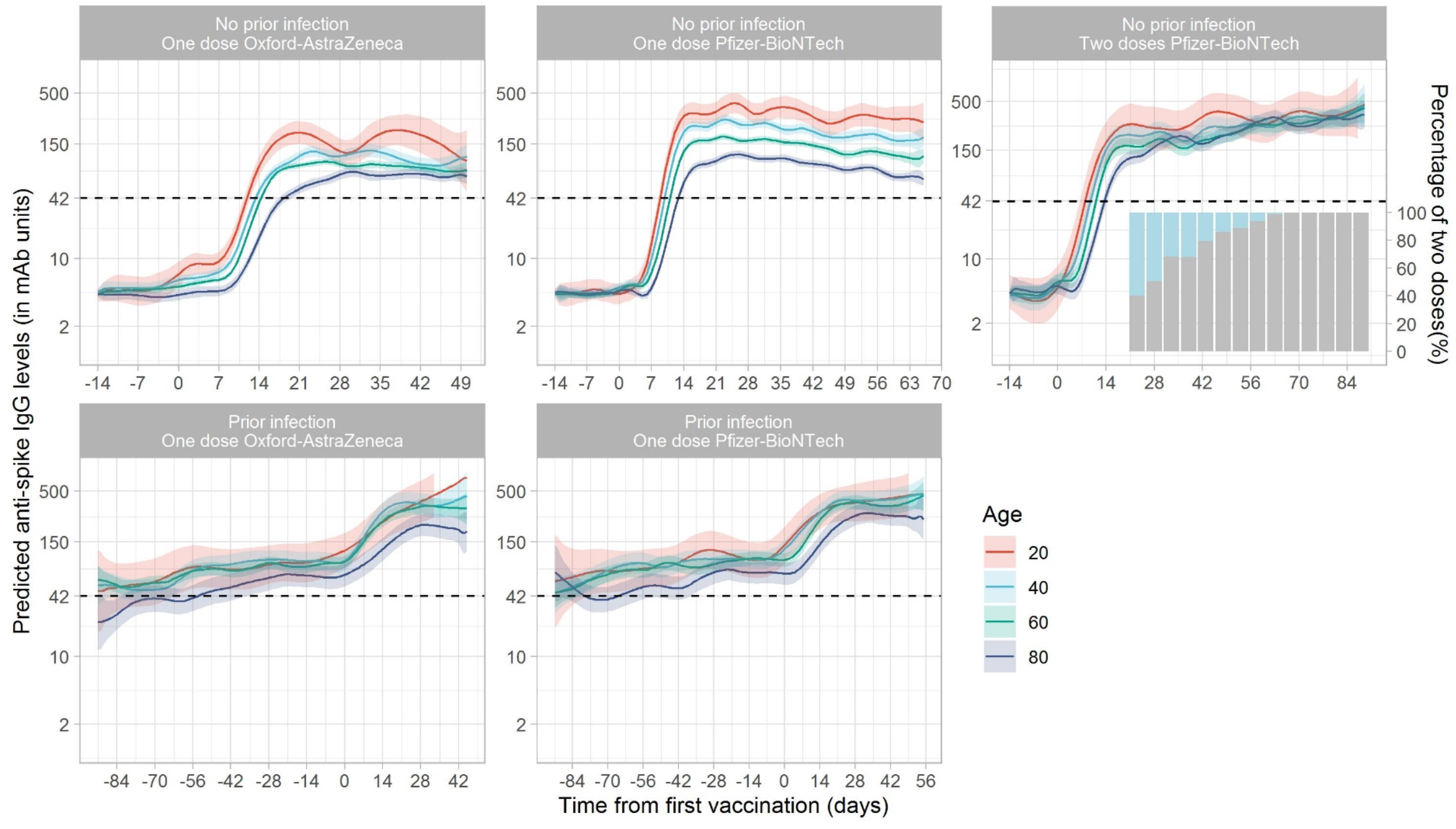
Predicted anti-spike IgG levels (in mAb units) by time from first vaccination, according to vaccine type and prior infection status. Predicted levels are plotted on a log scale. Black dotted line indicates the threshold of IgG positivity (42 units). Line colour indicates response predicted for ages 20, 40, 60, and 80 years (see **Figure S8** for all ages and **Figure 4** for comparisons of vaccine type by age). The bars in ‘Two doses Pfizer-BioNTech’ plot show the percentage of people having had two doses of vaccines by each timepoint (Gray: had two doses; Blue: had only one dose)

**Figure 4.**
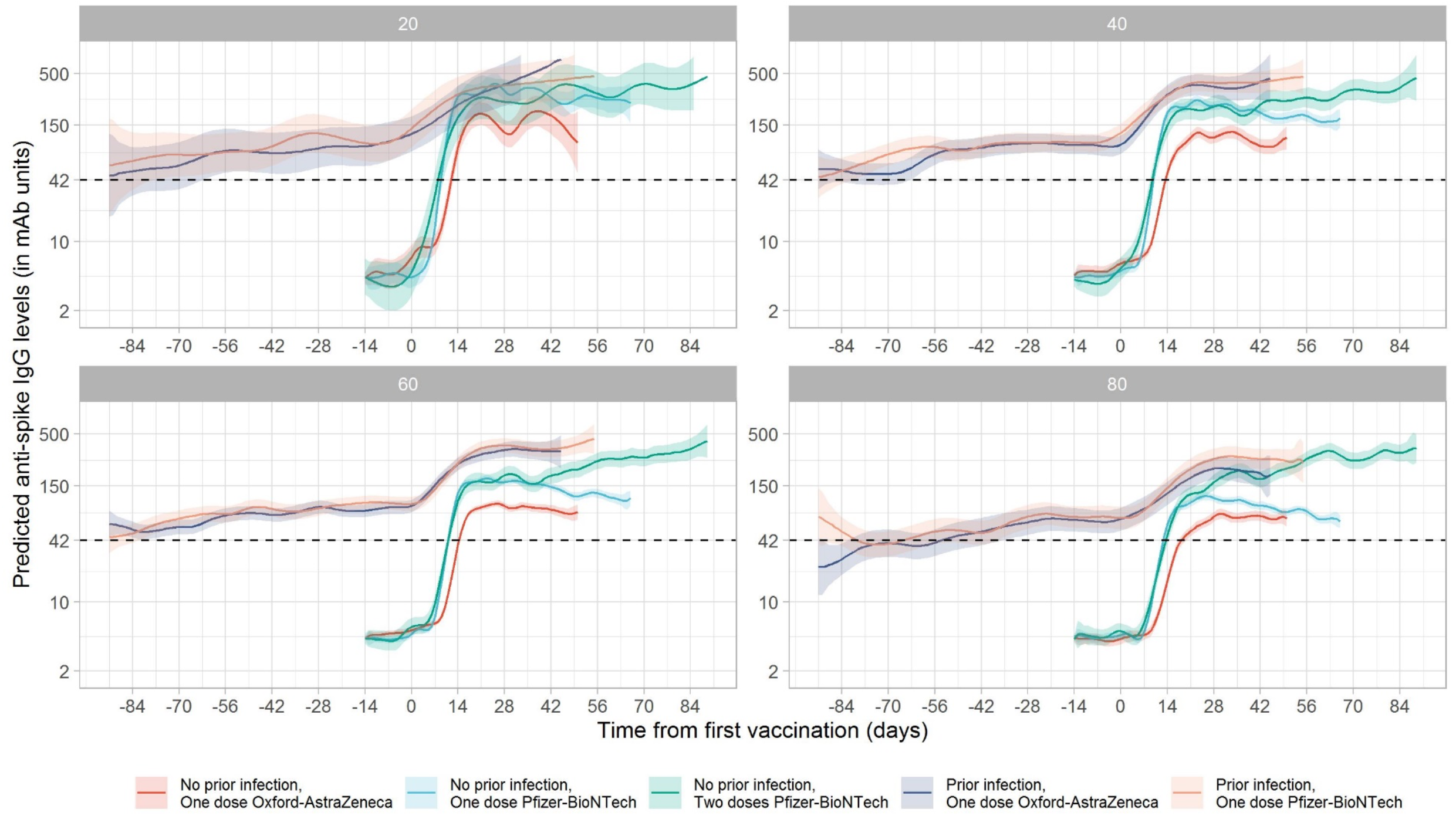
Predicted anti-spike IgG levels (in mAb units) by time from first vaccination for ages 20, 40, 60, and 80 years. (full models shown in **Figure S8**, plotted by vaccine in **Figure 3**). Line colour indicates predicted response for different vaccine type and prior infection status. Data identical to **Figure 3**, but **Figure 3** panels represent age rather than vaccine type.

For two doses of Pfizer-BioNTech vaccine, high anti-spike IgG levels were achieved 28 days after the first vaccination regardless of age, for example, 163 (95%CI 136-196), 192 (158-232), 232 (179-301), and 259 (153-440) ng/ml equivalents for 80-, 60-, 40-, and 20-year-olds (**Table S2**). The anti-spike IgG levels after receiving one dose of Pfizer-BioNTech compared with two doses were similar in younger ages but were substantially attenuated at older ages, with differences starting earlier post-first vaccine and attenuating more rapidly with increasing age (Figure 4).

In participants with evidence of prior infection, whilst vaccination increased antibody levels at all ages, the absolute increases were more modest. Participants of older ages with prior infection had lower IgG levels compared with younger ages both before and after vaccination (Figure 3). There was no evidence of a difference in response to vaccination after prior infection between those receiving Pfizer-BioNTech or Oxford-AstraZeneca vaccines (Figures 3, 4). At intermediate ages, antibody levels were significantly higher with a single dose following natural infection than two Pfizer-BioNTech doses, whereas two doses achieved similar antibody levels to one dose following natural infection at younger and older ages.

### Latent class analysis of antibody trajectory in participants without prior infection

We next investigated whether there was any evidence for a ‘non-responder’ group, particularly as means were only slightly above the positivity threshold for older individuals receiving the Oxford-AstraZeneca vaccine. Latent class mixed models identified four classes of antibody responses post-vaccination for both vaccine types (**Figure 5, Table S3**), with similar trajectories in each group for the two vaccines. In a ‘plausibly previously infected’ group (class 1, red line), estimated to comprise 5.9% of those receiving single-dose Oxford-AstraZeneca or Pfizer-BioNTech, participants’ anti-spike IgG levels started higher pre-vaccination (but below the threshold for positivity) and rose rapidly. In a ‘high response’ group (class 2, blue line), IgG levels increased rapidly and to a higher level before plateauing; 31.6% and 63.5% of participants who received single-dose Oxford-AstraZeneca and Pfizer-BioNTech respectively fell into this group. A medium response’ group (class 3, green line), with mean antibody levels slightly below the ‘high’ response group but still above the positivity threshold, comprised 58.7% and 27.5% of participants respectively. Lastly, participants in a ‘low response’ group (class 4, purple line) had mean IgG levels below the positivity threshold throughout, peaking at ∼10 mAb units, and their response was delayed. A similar percentage, 5.8% and 5.1% of participants receiving the Oxford-AstraZeneca and Pfizer-BioNTech vaccines, respectively, fell in this group. ‘Low response’ participants were older and ‘high response’ participants younger for both vaccines (**Figure S9**). ‘Low responders’ also had a higher proportion of males for Oxford-AstraZeneca vaccine and people with long-term health conditions for both vaccines (p<0.001).

**Figure 5.**
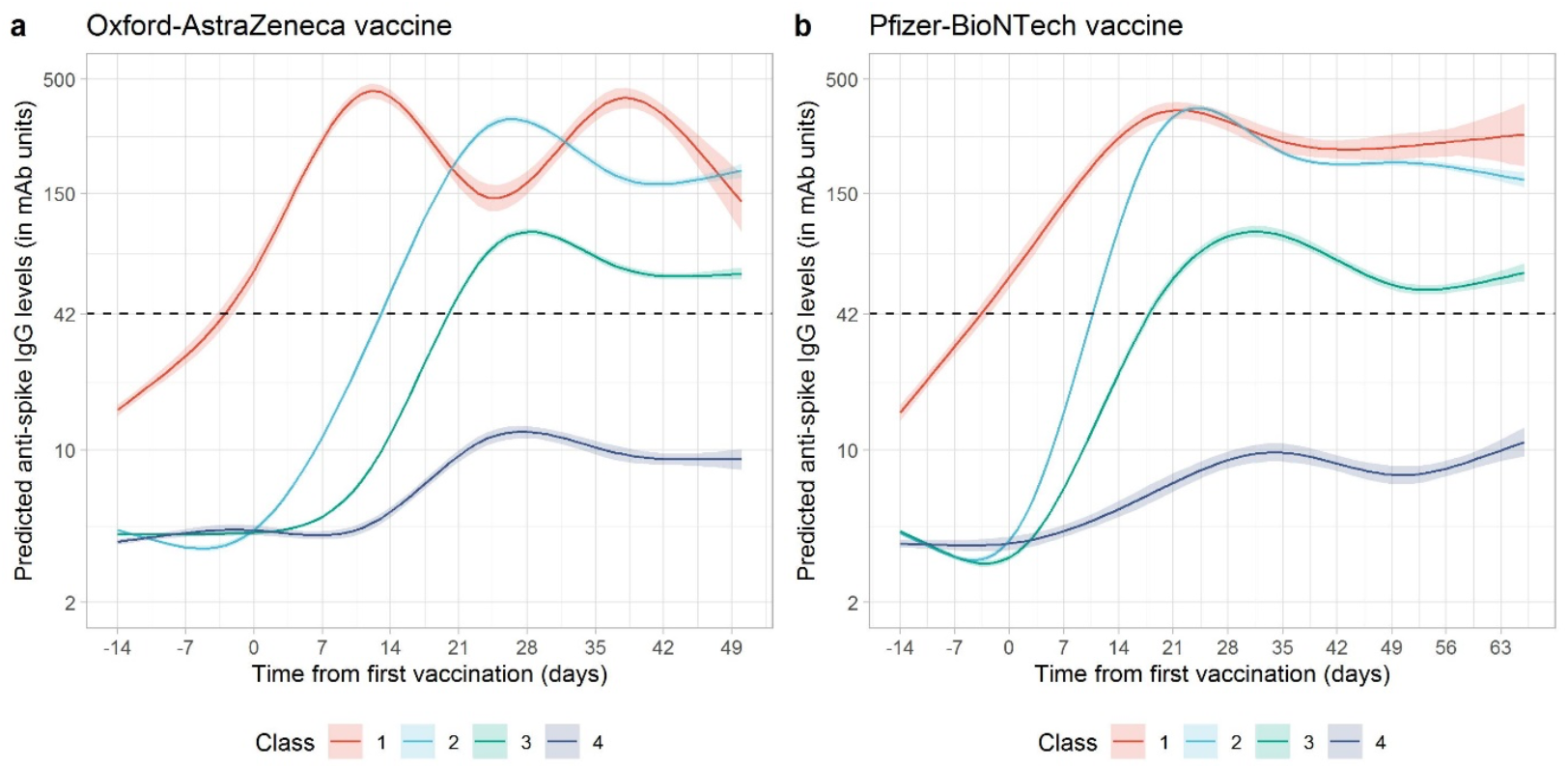
Predicted anti-spike IgG trajectory in participants without prior infection by class identified from latent class mixed models, using data from 14 days before vaccination to the 90^th^ percentile of the observed time points after vaccination. Panel A: One dose Oxford-AstraZeneca vaccine, no evidence of prior infection. Panel B: One dose Pfizer-BioNTech vaccine, no evidence of prior infection. Black dotted line indicates the threshold of IgG positivity (42 units). Distribution of factors by class membership shown in **Supplementary Table 3**. Class 1=’plausibly previously infected’ group (3.9% AZ, 3.9% Pfizer), 2=’high response’ group (31.6% AZ, 63.5% Pfizer), 3=’medium response’ group (58.7% AZ, 27.5% Pfizer), 4=’low response group’ (5.8% AZ, 5.1% Pfizer)

## Discussion

In this study based on 45,965 vaccinated participants from a large random sample of the UK population, we show post-vaccine anti-spike IgG responses vary by prior infection status, age, sex, the vaccine type, and number of doses received. In those who were previously infected, all age groups achieved high antibody response after first vaccination, in keeping with immune priming by natural infection followed by a vaccine boost. In those without evidence of prior infection, older participants had lower responses than younger participants after receiving a single dose of vaccine, with especially marked effects in those over 60 years; fewer older participants seroconverted, and quantitative antibody responses rose more slowly and to a lower level. Two vaccine doses in a conventional prime-boost regimen achieved high responses across all age groups, and particularly increased the number of older people seroconverting to similar levels to those receiving one dose after prior infection, as recently reported in a much smaller number of younger individuals^12^. Participants who received a single dose of Oxford-AstraZeneca vaccine had lower absolute antibody levels and their response was slower than those who received a single dose of Pfizer-BioNTech vaccine. However, the antibody levels in participants who received a single dose of Pfizer-BioNTech waned over time, whereas levels remained approximately constant after a single dose of Oxford-AstraZeneca. Importantly we did not identify any group who did not respond at all to vaccination, however, we did identify a non-negligible group (∼6%) of low responders to both vaccines.

The relative differences in vaccine response by participant demographics are similar to those reported by the Real-time Assessment of Community Transmission-2 (REACT-2) study which found a lower antibody positivity on a binary point-of-care lateral flow assay after a single dose of Pfizer vaccine with increasing age, and a consistently high response across all ages in people with prior infection^13^. However, our results showed a much higher antibody response than reported in REACT-2, especially in older people, despite being collected at similar times post vaccination and over similar calendar time in the UK. For example, REACT-2 reported seropositivity was 48.7% and 34.7% for people aged 70-79 and over 80 years 21 days after a single dose of Pfizer-BioNTech vaccine, while we found anti-spike IgG positivity was around 75-80%. These differences likely reflect the lower sensitivity of the assay used in REACT-2, despite efforts to adjust for this in the analysis^13^. The fact that we found mean quantitative responses were not far from the positivity threshold, particularly for older age groups, demonstrates the challenge in applying binary thresholds to what are essentially continuous data and supports differences in positivity rates between studies being attributable, at least in part, to varying, somewhat arbitrary, thresholds defining positivity. This is particularly important given the fact that antibody levels required for protection are still unclear, with at least one study using the same assay as our study identifying a gradient of protection associated with quantitative antibody levels below the positivity threshold following previous infection^14^. Our study provides additional comparative data on antibody responses following the Oxford-AstraZeneca vaccine. Studies in healthcare workers also support an inverse association between antibody response and age in those receiving a single dose of Pfizer-BioNTech^15,16^ or Oxford-AstraZeneca vaccine^6^.

We found that antibody positivity was also associated with sex. In those without evidence of previous infection, at older ages females had a higher probability of being IgG positive than males, and females were more likely to be in the ‘high response’ group from the latent class model. Sex differences in antibody levels have also been described following natural infection, e.g., a study of infected healthcare workers found that anti-S and neutralizing antibodies declined faster in males than in females^17^. These findings are consistent with observations that females generate stronger humoral immunity and greater vaccine efficacy than males^18,19^. However, a UK study on 3,610 healthcare workers did not find any association between sex and IgG positivity after a single dose of Oxford-AstraZeneca or Pfizer-BioNTech vaccine,^6^ possibly explained by our finding that sex differences in antibody responses become more marked as age increases over 60 years and older, and the median healthcare worker age was 41 years.

Consistent with several previous studies,^7,8,20^ we found that in previously infected participants, a single dose of Oxford-AstraZeneca or Pfizer-BioNTech vaccine led to high anti-spike IgG antibody positivity and quantitative anti-spike IgG levels. Given limited vaccine supplies, this supports prioritising those without evidence of previous infection for vaccination, and in particular delaying or even omitting second doses in those with robust serological evidence of previous infection. An additional finding from our study is that receiving two vaccine doses significantly increased seropositivity and antibody levels in older participants, but the incremental increase in 20-40 year-olds with second vaccine was much smaller, at least over the short-term. This suggests older age groups should be prioritised for second vaccination if supplies are limited. However, as discussed in more detail elsewhere^6^, there is an incomplete relationship between protection from infection and seroconversion, with rates of seroconversion post first dose vaccination exceeding the proportional reduction in symptomatic infection seen. Therefore, vaccine efficacy against clinical outcomes as well as antibody responses should contribute to prioritisation decisions.

Our latent class analysis identified four distinct type of vaccine responses, but interestingly no “complete non-responder” class. The response class with lower peak and delayed rises in IgG was more common in older participants and those with long term health conditions, but comprised a similar percentage receiving the different vaccines, suggesting a common biological cause. Further follow-up is needed to identify whether the modest increases in absolute levels achieved still lead to some degree of protection against key outcomes such as hospitalisation, death or onward transmission, and if not, whether a second vaccine dose (either using the same or different type of vaccine) substantially boost this initial sub-optimal response. This low-responder group could be identified by monitoring antibody titres as early as day 28, at which point titres below the assay threshold of 42 mAb units would be highly indicative of non-response. Similar underlying latent classes were identified following single doses of the two vaccines, with different mean responses overall due to different percentages estimated to fall into the ‘high’ and ‘medium’ response classes for Oxford-AstraZeneca and Pfizer-BioNTech vaccines; the impact of the second Oxford-AstraZeneca dose in such a general population cohort is yet unknown. Further studies are also required to assess whether different degrees of response are associated with different rates of waning over time and different levels of protection against clinical outcomes. In particular a recent study in 10,412 residents of long-term care facilities showed 65% and 68% protection against laboratory confirmed SARS-CoV-2 positivity 28-42 days after vaccination with the Oxford-AstraZeneca and Pfizer-BioNTech vaccines respectively, suggesting that any differences between vaccines in antibody responses may have limited impact on outcomes, at least in the short term^21^. Similar short-term (6 week) protection against symptomatic infection, hospitalisation and death with single doses of both vaccines was also seen in adults over 70 years in England^22^.

Limitations of our study include currently insufficient data to analyse responses following two doses of Oxford-AstraZeneca vaccine. Further follow up will be required to assess the duration of responses to all vaccines and how variation in interval between first and second doses impacts this. Although our study is broadly representative of those vaccinated to date in the UK, vaccination prioritisation means that we have fewer data on healthy younger adults. Although we show that long-term health conditions as a group are associated with lower antibody responses, additional studies of responses in specific conditions are required to understand its significance for vaccine protection. Our study assesses responses using a single assay; however, responses are calibrated to a monoclonal antibody such that these can be readily compared with other studies. Neutralising antibody and T cell responses were not assayed in this study. However, a recent much smaller study of T-cell responses in healthcare workers found qualitatively similar findings to our study in terms of responses to vaccination^23^.

In summary, in this population-representative study of individuals vaccinated to date in the UK, vaccination results in detectable SARS-CoV-2 anti-spike IgG in the majority of individuals after first vaccination. High rates of seroconversion and high quantitative antibody levels following one dose of vaccine after previous infection and in younger previously uninfected individuals potentially supports single dose or delayed second dose vaccination in these groups if vaccine supplies are limited, although the potential for this to lead to antigenic evolution requires investigation^24^. Further data from this study and others will be needed to assess the extent to which quantitative antibody levels can be used as a correlate of vaccine-mediated protection.

## Online Methods

### Population and data

We used data from the UK’s Office for National Statistics (ONS) COVID-19 Infection Survey (CIS) (ISRCTN21086382) from 26 April 2020 to 6 April 2021. The survey randomly selects private households on a continuous basis from address lists and previous surveys conducted by the ONS or the Northern Ireland Statistics and Research Agency to provide a representative sample across the four countries comprising the UK (England, Wales, Northern Ireland, Scotland). Following verbal agreement to participate, a study worker visited each household to take written informed consent. This consent was obtained from parents/carers for those 2-15 years, while those 10-15 years also provided written assent. Children aged <2 years were not eligible for the study. At the first visit, participants were asked for (optional) consent for follow-up visits every week for the next month, then monthly for 12 months from enrolment. For a random 10% of households, those ≥16 years were invited to provide blood monthly for serological testing from enrolment. Nose and throat swabs were taken from all consenting household members, according to the follow-up schedule they agreed to at enrolment. Any individual ≥16 years from a household where anyone tested positive on a nose and throat swab was also invited to provide blood at all subsequent monthly visits. Participants provided survey data on socio-demographic characteristics and vaccination status. Details on the sampling design are provided elsewhere^25^. The study protocol is available at https://www.ndm.ox.ac.uk/covid-19/covid-19-infection-survey/protocol-and-information-sheets. The study received ethical approval from the South Central Berkshire B Research Ethics Committee (20/SC/0195).

Vaccination data were reported by participants to the CIS and also obtained from linkage to the National Immunisation Management Service (NIMS), which holds a database of all individuals vaccinated in National Health Service COVID-19 vaccination programme in England. Similar linked administrative data was not available for Northern Ireland, Scotland, and Wales. Information on the date, doses, and type of vaccination were included in the dataset.

Only participants that received at least one dose of the Oxford-AstraZeneca or Pfizer-BioNTech vaccine were included; other vaccines were very rarely reported. Participants ≥16 years who had received at least one dose of vaccine from 8 December 2020 onwards with one or more antibody measurements from 91 days before their first vaccination date through to 6 April 2021 were included.

### Laboratory testing

SARS-CoV-2 antibody levels were measured using an ELISA detecting anti-trimeric spike IgG developed by the University of Oxford^25,26^. All testing was performed at the University of Oxford. Normalised results are reported in ng/ml of mAb45 monoclonal antibody equivalents. Up to 26 February 2021, the assay was performed using a fluorescence detection mechanism as previously described, using a threshold of 8 million units to identify positive samples.^26^ Subsequent testing was performed with a CE-marked version of the assay, the Thermo Fisher OmniPATH 384 Combi SARS-CoV-2 IgG ELISA, which uses the same antigen, with a colorimetric detection system. mAb45 is the manufacturer-provided monoclonal antibody calibrant for this quantitative assay. To allow conversion of fluorometrically determined values in arbitrary units, 3840 samples were run in parallel on both systems and compared. A piece-wise linear regression was used to generate the following conversion formula:

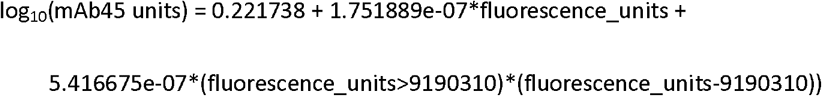

A threshold of ≥42 ng/ml was used to identify IgG-positive samples, corresponding to the 8 million units with fluorescence detection. In this analyses, measurements <2 ng/ml (395 observations, 0.4%) and >500 ng/ml (7,707 observations, 7%) were truncated at 2 and 500 ng/ml, respectively.

PCR of combined nose and throat swabs was undertaken using the Thermo Fisher TaqPath SARS-CoV-2 assay at high-throughput national “Lighthouse” laboratories in Glasgow and Milton Keynes (to 8 February 2021). PCR outputs were analysed using UgenTec FastFinder 3.300.5, with an assay-specific algorithm and decision mechanism that allows conversion of amplification assay raw data into test results with minimal manual intervention. Samples are called positive if at least a single N-gene and/or ORF1ab are detected (although S-gene cycle threshold (Ct) values are determined, S-gene detection alone is not considered sufficient to call a sample positive^25^) and PCR traces exhibit an appropriate morphology.

### Statistical analysis

Participants with a SARS-CoV-2 PCR-positive nose/throat swab or a prior positive anti-spike IgG antibody result at any time prior to vaccination were considered to have been previously infected with SARS-CoV-2, irrespective of whether they had reported previous symptoms or not. Regular PCR results from the survey were included in this classification, but not self-reported PCR or lateral flow test results obtained outside the study. We used multivariable logistic and linear generalized additive models (GAMs) to investigate binary (positive/negative) and quantitative (log_10_(mAb45 units)) anti-spike IgG antibody measurements post-first vaccination. Given the prior hypothesis that response to vaccination would vary differentially by age and time according to vaccine type and prior infection, we built separate models by vaccine type, for those receiving one or two vaccinations, and by prior infection status. For participants receiving one vaccine dose, four models were fitted, for each vaccine and in those with and without evidence of prior infection. Two dose models were only fitted for those receiving Pfizer-BioNTech vaccine without evidence of prior infection, as sample sizes were small for other groups.

Models were adjusted for participant age using a tensor product of B-splines to allow for non-linearity and interaction between age and time since vaccination. The smoothing penalty was selected using fast restricted maximum likelihood (fREML) as implemented in the mcgv R package. We included a random intercept for each participant to account for repeated measurements using a random effect smoother with the number of basis functions equal to the number of participants. The date of the first vaccination was set as t=0. For those with no prior evidence of infection, we truncated time at t=0 and t=-14 for logistic and linear models respectively (t=-14 for linear models to estimate IgG baseline pre-vaccination). We excluded measurements taken after the 90^th^ percentile of observed time points for all models to avoid undue influence from outliers at late time points. Any participant receiving a second Pfizer-BioNTech dose after the 90^th^ percentile for the single Pfizer-BioNTech dose group (61 days) was censored at this timepoint and included in the one dose group (1,383 [3%] participants were censored in this way). Age was truncated at 85 years in all analyses to avoid outlier influence.

To investigate predictors of antibody response in those without prior evidence of infection, we considered the latest antibody measurement per participant between 14-60 days post first vaccine. We used multivariable logistic regression to examine the association between antibody positivity and vaccine type and doses received by this measurement time, demographic factors (age, sex, ethnicity), household size, deprivation ranking (index of multiple deprivation (IMD) in England and equivalent percentile ranking in Wales, Northern Ireland and Scotland), whether the participant reported working in patient-facing healthcare or social care, whether they reported working in a care home (any role), and whether they reported having a long-term health condition. Non-linearity in age was accounted for using restricted natural cubic splines with internal knots at the 20^th^, 40^th^, 60^th^, and 80^th^ percentiles of unique values, and boundary knots at 5^th^ and 95^th^ percentiles. We tested for and added interactions between age and other variables if the interaction p-value was <0.05.

For those without evidence of prior infection who received a single dose of vaccine we also investigated whether we could identify distinct patterns of antibody responses, using latent class mixed models (LCMM) to identify sub-populations with different antibody trajectories after first vaccination. Natural cubic splines (internal knots at the 20^th^, 40^th^, 60^th^, 80^th^ percentiles of unique values, and boundary knots at 5^th^ and 95^th^ percentiles) were used to model time since vaccination as a fixed effect and a random intercept was added to account for individual variability. Within-class between-individual heterogeneity may also be present in the trajectories; however, models accounting for random slopes failed to converge. Age with natural cubic splines (same as above), sex, reported long-term health conditions, and whether the participant was a healthcare worker were included as covariates for class membership. The number of classes was determined by minimizing the Bayesian information criterion (BIC) for each vaccine, and then fitting the maximum number of classes (three) to both groups for comparability.

Analyses were performed using the mgcv, splines, and lcmm libraries in R (v.3.6).

## Data Availability

Data are still being collected for the COVID-19 Infection Survey. De-identified study data are available for access by accredited researchers in the ONS Secure Research Service (SRS) for accredited research purposes under part 5, chapter 5 of the Digital Economy Act 2017. For further information about accreditation, contact or visit the SRS website.

## Acknowledgements

We are grateful for the support of all COVID-19 infection survey participants.

**Office for National Statistics:** Sir Ian Diamond, Iain Bell, Emma Rourke, Ruth Studley, Alex Lambert, Tina Thomas.

**Office for National Statistics COVID Infection Survey Analysis and Operations teams**, in particular Daniel Ayoubkhani, Russell Black, Antonio Felton, Megan Crees, Joel Jones, Lina Lloyd, Esther Sutherland. University of Oxford, Nuffield Department of Medicine: Ann Sarah Walker, Derrick Crook, Philippa C Matthews, Tim Peto, Emma Pritchard, Nicole Stoesser, Karina-Doris Vihta, Jia Wei, Alison Howarth, George Doherty, James Kavanagh, Kevin K Chau, Stephanie B Hatch, Daniel Ebner, Lucas Martins Ferreira, Thomas Christott, Brian D Marsden, Wanwisa Dejnirattisai, Juthathip Mongkolsapaya, Sarah Cameron, Phoebe Tamblin-Hopper, Magda Wolna, Rachael Brown, Sarah Hoosdally, Richard Cornall, David I Stuart, Gavin Screaton.

**University of Oxford, Nuffield Department of Population Health:** Koen Pouwels.

**University of Oxford, Big Data Institute:** David W Eyre, Katrina Lythgoe, David Bonsall, Tanya Golubchik, Helen Fryer.

**University of Oxford, Radcliffe Department of Medicine:** John Bell.

**Oxford University Hospitals NHS Foundation Trust:** Stuart Cox, Kevin Paddon, Tim James.

**University of Manchester:** Thomas House.

**Public Health England:** John Newton, Julie Robotham, Paul Birrell.

**IQVIA:** Helena Jordan, Tim Sheppard, Graham Athey, Dan Moody, Leigh Curry, Pamela Brereton.

**National Biocentre:** Ian Jarvis, Anna Godsmark, George Morris, Bobby Mallick, Phil Eeles.

**Glasgow Lighthouse Laboratory:** Jodie Hay, Harper VanSteenhouse.

**Department of Health:** Jessica Lee.

## Author Contributions

The study was designed and planned by ASW, JF, JB, JN, IB, ID and KBP, and is being conducted by ASW, IB, RS, ER, AH, BM, TEAP, PCM, NS, SH, EYJ, DIS, DWC and DWE. This specific analysis was designed by JW, DWE, ASW, and KBP. JW and KBP contributed to the statistical analysis of the survey data. JW, DWE, KBP and ASW drafted the manuscript and all authors contributed to interpretation of the data and results and revised the manuscript. All authors approved the final version of the manuscript.

