## Supplementary material for "The impact of SARS-CoV-2 vaccines on antibody responses in the general population in the United Kingdom"

### Supplementary tables

| | No evidence of prior infection | | | Evidence of prior infection | | | |
|---|---|---|---|---|---|---|---|
| | One dose AZ (N=23368) | One dose Pf (N=14894) | Two doses Pf(N=1869) | One dose AZ (N=3767) | One dose Pf (N=2067) | Total (N=45965) | p value |
| **Duration between two doses [Median (IQR)]** | | | 31 (21-47) | | | | |
| **Age** | | | | | | | < 0.001 |
| **Median** | 63 | 66 | 70 | 61 | 62 | 64 | |
| **IQR** | 55, 70 | 54, 73 | 53, 80 | 52, 68 | 50, 70 | 54, 71 | |
| **Sex** | | | | | | | < 0.001 |
| **Female** | 12510 (53.5%) | 8497 (57.0%) | 1153 (61.7%) | 2034 (54.0%) | 1136 (55.0%) | 25330 (55.1%) | |
| **Male** | 10858 (46.5%) | 6397 (43.0%) | 716 (38.3%) | 1733 (46.0%) | 931 (45.0%) | 20635 (44.9%) | |
| **Ethnicity** | | | | | | | < 0.001 |
| **Non-white** | 1110 (4.8%) | 817 (5.5%) | 118 (6.3%) | 241 (6.4%) | 148 (7.2%) | 2434 (5.3%) | |
| **White** | 22258 (95.2%) | 14077 (94.5%) | 1751 (93.7%) | 3526 (93.6%) | 1919 (92.8%) | 43531 (94.7%) | |
| **Household size** | | | | | | | < 0.001 |
| **1** | 5019 (21.5%) | 3302 (22.2%) | 501 (26.8%) | 721 (19.1%) | 395 (19.1%) | 9938 (21.6%) | |
| **2** | 12990 (55.6%) | 8260 (55.5%) | 985 (52.7%) | 1916 (50.9%) | 1072 (51.9%) | 25223 (54.9%) | |
| **3** | 2863 (12.3%) | 1688 (11.3%) | 168 (9.0%) | 545 (14.5%) | 312 (15.1%) | 5576 (12.1%) | |
| **4** | 1797 (7.7%) | 1130 (7.6%) | 145 (7.8%) | 403 (10.7%) | 202 (9.8%) | 3677 (8.0%) | |
| **5+** | 699 (3.0%) | 514 (3.5%) | 70 (3.7%) | 182 (4.8%) | 86 (4.2%) | 1551 (3.4%) | |
| **Deprivation percentile** | | | | | | | < 0.001 |
| **Median** | 63 | 64 | 64 | 60 | 57 | 63 | |
| **IQR** | 40, 82 | 40, 83 | 38, 83 | 35, 81 | 32, 79 | 39, 82 | |
| **Report working in patient facing healthcare** | | | | | | | < 0.001 |
| **No** | 22860 (97.8%) | 13430 (90.2%) | 1466 (78.4%) | 3636 (96.5%) | 1828 (88.4%) | 43220 (94.0%) | |
| **Yes** | 508 (2.2%) | 1464 (9.8%) | 403 (21.6%) | 131 (3.5%) | 239 (11.6%) | 2745 (6.0%) | |
| **Report working in person facing social care** | | | | | | | < 0.001 |
| **No** | 23149 (99.1%) | 14548 (97.7%) | 1800 (96.3%) | 3717 (98.7%) | 2014 (97.4%) | 45228 (98.4%) | |
| **Yes** | 219 (0.9%) | 346 (2.3%) | 69 (3.7%) | 50 (1.3%) | 53 (2.6%) | 737 (1.6%) | |
| **Report working in a care home (any role)** | | | | | | | < 0.001 |
| **No** | 23195 (99.3%) | 14551 (97.7%) | 1778 (95.1%) | 3701 (98.2%) | 2011 (97.3%) | 45236 (98.4%) | |
| **Yes** | 173 (0.7%) | 343 (2.3%) | 91 (4.9%) | 66 (1.8%) | 56 (2.7%) | 729 (1.6%) | |
| **Report having long-term health condition** | | | | | | | < 0.001 |
| **No** | 15774 (67.5%) | 9597 (64.4%) | 1217 (65.1%) | 2645 (70.2%) | 1398 (67.6%) | 30631 (66.6%) | |
| **Yes** | 7594 (32.5%) | 5297 (35.6%) | 652 (34.9%) | 1122 (29.8%) | 669 (32.4%) | 15334 (33.4%) | |

**Table S1. Participants' characteristics by cohort.** AZ: Oxford-AstraZeneca vaccine. Pf: Pfizer-BioNTech vaccine. Note: higher deprivation percentile means living in a less deprived area.

| | | No prior infection | | | Prior infection | |
|---|---|---|---|---|---|---|
| **Time (days)** | **Age** | **AZ one dose** | **Pfizer one dose** | **Pfizer two doses** | **AZ one dose** | **Pfizer one dose** |
| **Logistic model** | | Predicted probability of anti-spike IgG positivity (95%CI) | | | | |
| **0** | 20 | 1 (0-3) | 0 (0-1) | 5 (1-16) | 90 (82-95) | 94 (88-97) |
| **0** | 40 | 1 (0-1) | 0 (0-1) | 2 (1-5) | 85 (80-88) | 89 (84-92) |
| **0** | 60 | 0 (0-0) | 0 (0-0) | 1 (1-2) | 78 (75-82) | 81 (77-84) |
| **0** | 80 | 0 (0-0) | 0 (0-0) | 0 (0-1) | 70 (61-78) | 69 (59-77) |
| **7** | 20 | 9 (4-20) | 29 (17-45) | 60 (34-81) | 91 (83-96) | 97 (94-99) |
| **7** | 40 | 6 (4-10) | 13 (9-18) | 36 (23-52) | 88 (83-91) | 94 (90-96) |
| **7** | 60 | 5 (4-6) | 6 (4-8) | 18 (12-25) | 82 (78-85) | 87 (83-90) |
| **7** | 80 | 2 (1-4) | 3 (2-5) | 8 (4-14) | 74 (66-81) | 73 (64-81) |
| **14** | 20 | 79 (62-90) | 97 (92-99) | 95 (87-98) | 93 (86-97) | 99 (97-100) |
| **14** | 40 | 63 (55-71) | 95 (92-97) | 89 (80-94) | 92 (89-94) | 97 (95-99) |
| **14** | 60 | 49 (44-53) | 89 (86-91) | 78 (70-83) | 89 (87-92) | 95 (93-96) |
| **14** | 80 | 20 (14-28) | 66 (59-72) | 60 (49-70) | 83 (76-88) | 90 (84-93) |
| **21** | 20 | 90 (77-96) | 98 (94-99) | 98 (93-99) | 96 (89-98) | 99 (97-100) |
| **21** | 40 | 85 (78-90) | 96 (94-98) | 96 (91-98) | 95 (92-97) | 99 (97-99) |
| **21** | 60 | 81 (77-84) | 93 (91-95) | 91 (87-94) | 95 (93-96) | 98 (97-98) |
| **21** | 80 | 60 (52-68) | 81 (75-85) | 84 (77-89) | 90 (85-93) | 96 (93-98) |
| **28** | 20 | 90 (74-96) | 97 (93-99) | 97 (89-99) | 97 (92-99) | 99 (97-100) |
| **28** | 40 | 84 (76-89) | 95 (92-97) | 95 (90-98) | 97 (94-98) | 99 (98-100) |
| **28** | 60 | 79 (75-83) | 91 (89-93) | 93 (89-95) | 96 (95-97) | 98 (97-99) |
| **28** | 80 | 74 (66-80) | 85 (80-89) | 89 (84-93) | 94 (90-96) | 98 (95-99) |
| **35** | 20 | 94 (79-99) | 98 (95-99) | 93 (83-98) | 99 (93-100) | 99 (97-100) |
| **35** | 40 | 86 (78-91) | 96 (93-97) | 94 (88-97) | 98 (95-99) | 99 (98-100) |
| **35** | 60 | 80 (75-84) | 91 (89-93) | 94 (91-96) | 97 (96-98) | 98 (97-99) |
| **35** | 80 | 74 (67-80) | 81 (76-85) | 94 (90-97) | 95 (92-97) | 97 (95-99) |
| **42** | 20 | 92 (72-98) | 98 (94-99) | 95 (86-98) | 99 (93-100) | 100 (98-100) |
| **42** | 40 | 84 (75-90) | 96 (93-97) | 94 (88-97) | 99 (95-100) | 99 (98-100) |
| **42** | 60 | 79 (73-84) | 91 (88-93) | 93 (89-95) | 97 (95-99) | 98 (97-99) |
| **42** | 80 | 75 (67-81) | 82 (77-86) | 91 (85-95) | 96 (91-98) | 96 (93-98) |
| **49** | 20 | 80 (39-96) | 98 (94-99) | 95 (87-98) | - | 100 (98-100) |
| **49** | 40 | 84 (71-92) | 95 (92-97) | 94 (89-97) | - | 100 (97-100) |
| **49** | 60 | 78 (67-86) | 89 (85-92) | 93 (89-95) | - | 98 (96-99) |
| **49** | 80 | 78 (67-86) | 79 (74-84) | 91 (85-94) | - | 92 (86-96) |
| **Linear model** | | Predicted anti-spike IgG levels in mAb units (95%CI) | | | | |
| **0** | 20 | 7 (5-9) | 4 (3-6) | 5 (3-8) | 123 (82-185) | 139 (90-216) |
| **0** | 40 | 6 (5-7) | 5 (5-6) | 5 (4-7) | 96 (78-119) | 126 (102-156) |
| **0** | 60 | 5 (5-6) | 5 (4-5) | 6 (5-7) | 94 (81-108) | 98 (83-115) |
| **0** | 80 | 4 (4-5) | 5 (4-5) | 5 (4-6) | 69 (53-90) | 71 (55-92) |
| **7** | 20 | 9 (7-12) | 14 (11-18) | 30 (17-52) | 167 (111-251) | 220 (141-342) |
| **7** | 40 | 7 (7-8) | 10 (9-11) | 17 (13-22) | 169 (136-210) | 202 (162-250) |
| **7** | 60 | 6 (6-6) | 7 (7-8) | 10 (8-12) | 152 (132-176) | 138 (117-163) |
| **7** | 80 | 5 (4-5) | 5 (5-5) | 6 (5-8) | 88 (67-117) | 85 (66-108) |

| | | | | | | |
|---|---|---|---|---|---|---|
| **14** | 20 | 90 (71-115) | 293 (228-375) | 182 (110-301) | 242 (155-376) | 307 (204-464) |
| **14** | 40 | 50 (45-56) | 201 (181-225) | 151 (117-196) | 306 (243-386) | 301 (244-371) |
| **14** | 60 | 38 (36-40) | 127 (119-137) | 105 (86-128) | 248 (214-287) | 254 (217-298) |
| **14** | 80 | 16 (15-19) | 61 (56-67) | 48 (39-58) | 126 (96-166) | 143 (113-181) |
| **21** | 20 | 197 (152-257) | 325 (258-409) | 287 (173-478) | 318 (197-514) | 363 (243-542) |
| **21** | 40 | 113 (100-127) | 250 (225-277) | 219 (171-280) | 381 (299-485) | 398 (324-489) |
| **21** | 60 | 91 (85-97) | 173 (162-185) | 166 (136-202) | 312 (267-365) | 361 (307-424) |
| **21** | 80 | 52 (47-58) | 101 (93-110) | 123 (103-147) | 182 (141-236) | 223 (174-286) |
| **28** | 20 | 127 (94-171) | 334 (266-420) | 259 (153-440) | 389 (237-639) | 380 (245-591) |
| **28** | 40 | 113 (99-129) | 236 (214-261) | 232 (179-301) | 369 (286-477) | 402 (324-497) |
| **28** | 60 | 94 (87-100) | 163 (153-175) | 192 (158-232) | 347 (291-413) | 385 (328-452) |
| **28** | 80 | 73 (65-81) | 113 (104-123) | 163 (136-196) | 224 (173-290) | 281 (221-356) |
| **35** | 20 | 192 (135-274) | 359 (278-464) | 250 (157-400) | 485 (274-860) | 401 (250-644) |
| **35** | 40 | 125 (108-144) | 216 (193-242) | 196 (157-246) | 356 (267-476) | 404 (321-508) |
| **35** | 60 | 92 (85-100) | 159 (148-171) | 160 (133-193) | 345 (283-420) | 361 (305-429) |
| **35** | 80 | 71 (64-79) | 108 (99-117) | 212 (180-251) | 218 (165-288) | 295 (234-372) |
| **42** | 20 | 188 (123-288) | 287 (221-373) | 344 (215-551) | 622 (251-1542) | 424 (262-685) |
| **42** | 40 | 93 (80-109) | 202 (178-229) | 244 (192-310) | 412 (273-621) | 411 (320-527) |
| **42** | 60 | 86 (78-95) | 146 (134-159) | 191 (159-229) | 335 (259-433) | 353 (292-426) |
| **42** | 80 | 74 (66-83) | 94 (86-104) | 175 (145-211) | 200 (146-275) | 280 (222-353) |
| **49** | 20 | 107 (59-196) | 269 (206-351) | 382 (241-608) | - | 450 (260-777) |
| **49** | 40 | 108 (85-139) | 177 (155-202) | 264 (210-332) | - | 441 (336-579) |
| **49** | 60 | 79 (66-94) | 117 (106-129) | 218 (181-263) | - | 385 (312-475) |
| **49** | 80 | 73 (62-87) | 81 (74-89) | 220 (184-262) | - | 273 (212-351) |

**Table S2. Predicted probability of anti-spike IgG seropositivity and anti-spike IgG levels (in mAb units) with 95% confidence interval (CI) according to age, vaccine type and dose, and prior infection status by weeks after vaccination.** AZ: Oxford-AstraZeneca vaccine. Pf: Pfizer-BioNTech vaccine.

| | Oxford-AstraZeneca | | | | | Pfizer-BioNTech | | | | |
|---|---|---|---|---|---|---|---|---|---|---|
| | **Class 1 (N=867)** | **Class 2 (N=7097)** | **Class 3 (N=13163)** | **Class 4 (N=1297)** | **p value** | **Class 1 (N=547)** | **Class 2 (N=8951)** | **Class 3 (N=3876)** | **Class 4 (N=720)** | **p value** |
| **Percentage** | 3.9% | 31.6% | 58.7% | 5.8% | | 3.9% | 63.5% | 27.5% | 5.1% | |
| **Age** | | | | | < 0.001 | | | | | < 0.001 |
| **Median** | 58 | 60 | 64 | 68 | | 64 | 62 | 72 | 71 | |
| **IQR** | 50, 66 | 48, 68 | 57, 70 | 60, 73 | | 50, 73 | 49, 70 | 66, 77 | 62, 78 | |
| **Age group** | | | | | < 0.001 | | | | | < 0.001 |
| **16-34** | 73 (8.4%) | 709 (10.0%) | 167 (1.3%) | 22 (1.7%) | | 63 (11.5%) | 760 (8.5%) | 12 (0.3%) | 26 (3.6%) | |
| **35-54** | 274 (31.6%) | 1863 (26.3%) | 2412 (18.3%) | 165 (12.7%) | | 119 (21.8%) | 2240 (25.0%) | 231 (6.0%) | 76 (10.6%) | |
| **55-74** | 461 (53.2%) | 4119 (58.0%) | 9185 (69.8%) | 862 (66.5%) | | 250 (45.7%) | 4863 (54.3%) | 2209 (57.0%) | 354 (49.2%) | |
| **>75** | 59 (6.8%) | 406 (5.7%) | 1399 (10.6%) | 248 (19.1%) | | 115 (21.0%) | 1088 (12.2%) | 1424 (36.7%) | 264 (36.7%) | |
| **Sex** | | | | | < 0.001 | | | | | < 0.001 |
| **Female** | 453 (52.2%) | 4467 (62.9%) | 6505 (49.4%) | 600 (46.3%) | | 305 (55.8%) | 5577 (62.3%) | 1721 (44.4%) | 364 (50.6%) | |
| **Male** | 414 (47.8%) | 2630 (37.1%) | 6658 (50.6%) | 697 (53.7%) | | 242 (44.2%) | 3374 (37.7%) | 2155 (55.6%) | 356 (49.4%) | |
| **Report working in patient facing healthcare** | | | | | < 0.001 | | | | | < 0.001 |
| **No** | 834 (96.2%) | 6902 (97.3%) | 12966 (98.5%) | 1276 (98.4%) | | 475 (86.8%) | 7976 (89.1%) | 3791 (97.8%) | 692 (96.1%) | |
| **Yes** | 33 (3.8%) | 195 (2.7%) | 197 (1.5%) | 21 (1.6%) | | 72 (13.2%) | 975 (10.9%) | 85 (2.2%) | 28 (3.9%) | |
| **Report having long-term health condition** | | | | | < 0.001 | | | | | < 0.001 |
| **No** | 592 (68.3%) | 5104 (71.9%) | 8859 (67.3%) | 636 (49.0%) | | 355 (64.9%) | 6219 (69.5%) | 2096 (54.1%) | 344 (47.8%) | |
| **Yes** | 275 (31.7%) | 1993 (28.1%) | 4304 (32.7%) | 661 (51.0%) | | 192 (35.1%) | 2732 (30.5%) | 1780 (45.9%) | 376 (52.2%) | |
| **Membership probability (%)** | | | | | < 0.001 | | | | | < 0.001 |
| **Median** | 98 | 82 | 71 | 99 | | 94 | 89 | 86 | 100 | |
| **IQR** | 78, 100 | 61, 97 | 54, 92 | 84, 100 | | 75, 100 | 74, 98 | 64, 97 | 91, 100 | |

**Table S3. Characteristics of classes identified from latent class mixed models for single dose Oxford-AstraZeneca and Pfizer-BioNTech vaccines in those without evidence of prior infection.** Class 1='plausibly previously infected' group, 2='high response' group, 3='medium response' group, 4='low response group'.

### Supplementary figures

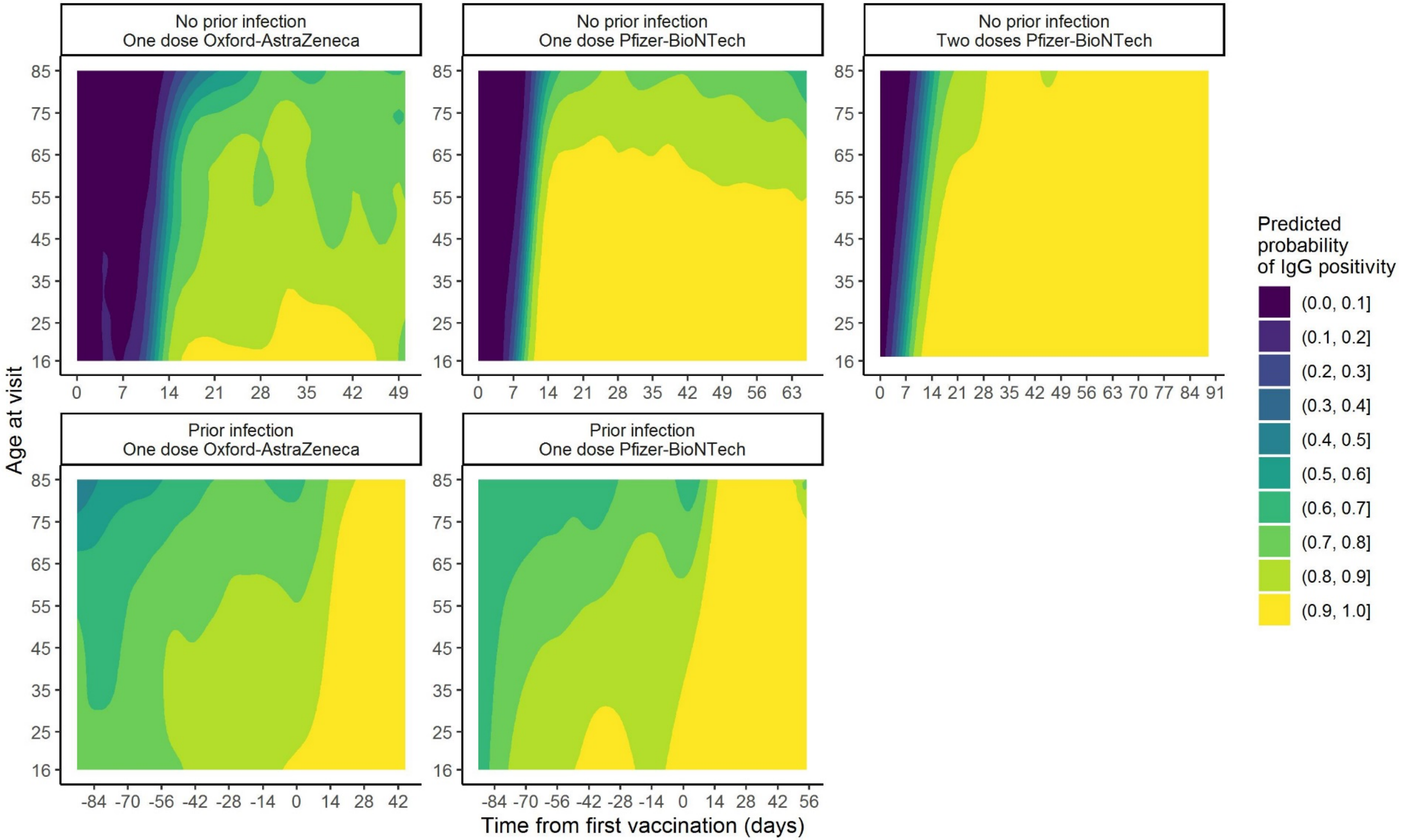


**Figure S1. Predicted probability of anti-spike IgG positivity by time from first vaccination and age, according to vaccine type and prior infection status (full model).** Predictions shown for specific ages in **Figure 1**. Observed data in **Supplementary Figures S3-S7**.

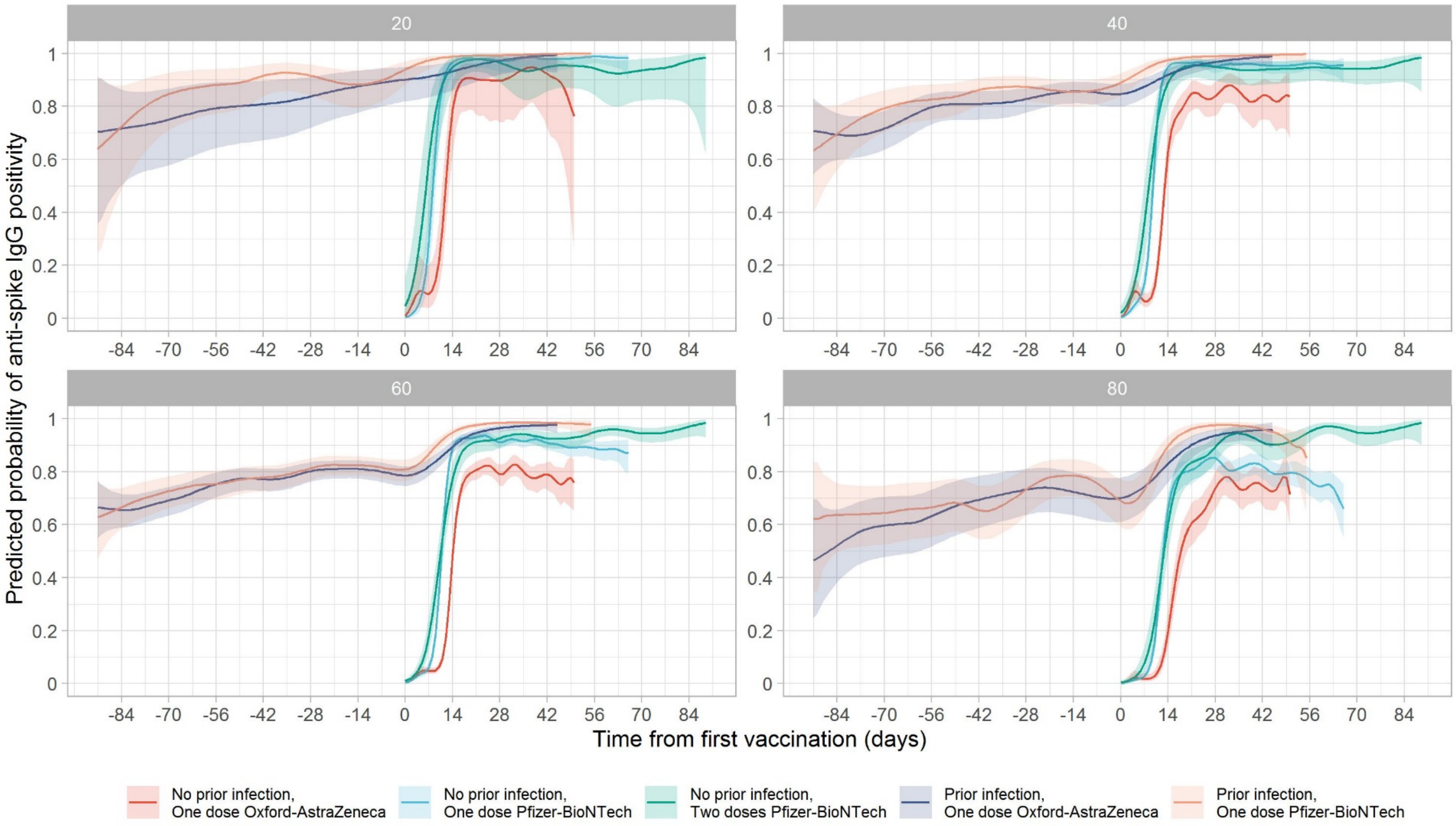


**Figure S2. Predicted probability of anti-spike IgG positivity by time from first vaccination for ages 20, 40, 60, and 80 years** (full models shown in **Figure S1**, plotted by vaccine in **Figure 1**)**.** Line colour indicates different vaccine type and prior infection status. Data identical to **Figure 1**, but Figure 1 panels represent age rather than vaccine type as here.

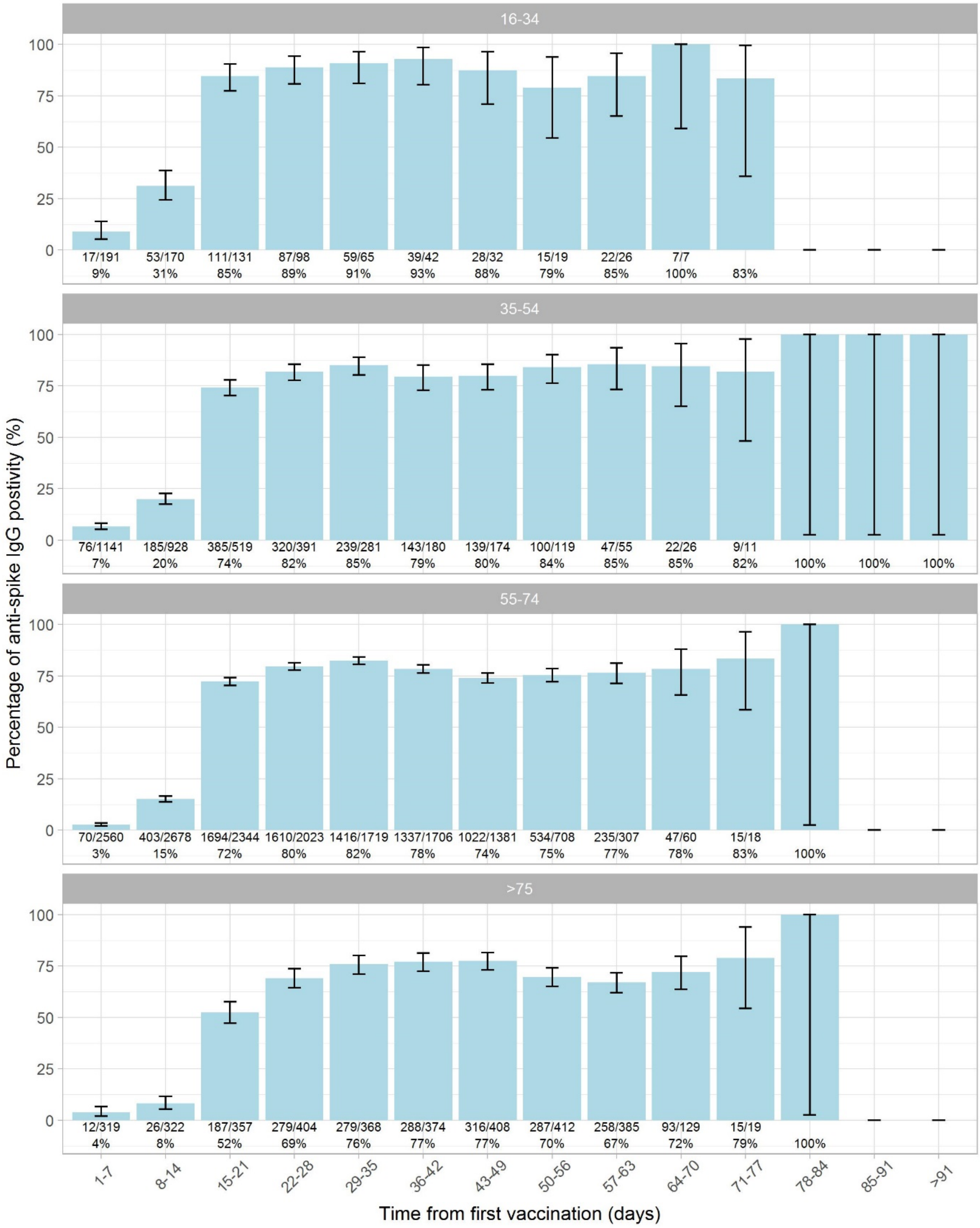


**Figure S3. Percentage (95% CI) anti-spike IgG positive by days after first vaccination in participants receiving a single dose of Oxford-AstraZeneca vaccine and without evidence of prior infection.** Results were divided into four age groups: 16-34, 35-54, 55-74, and >75 years.

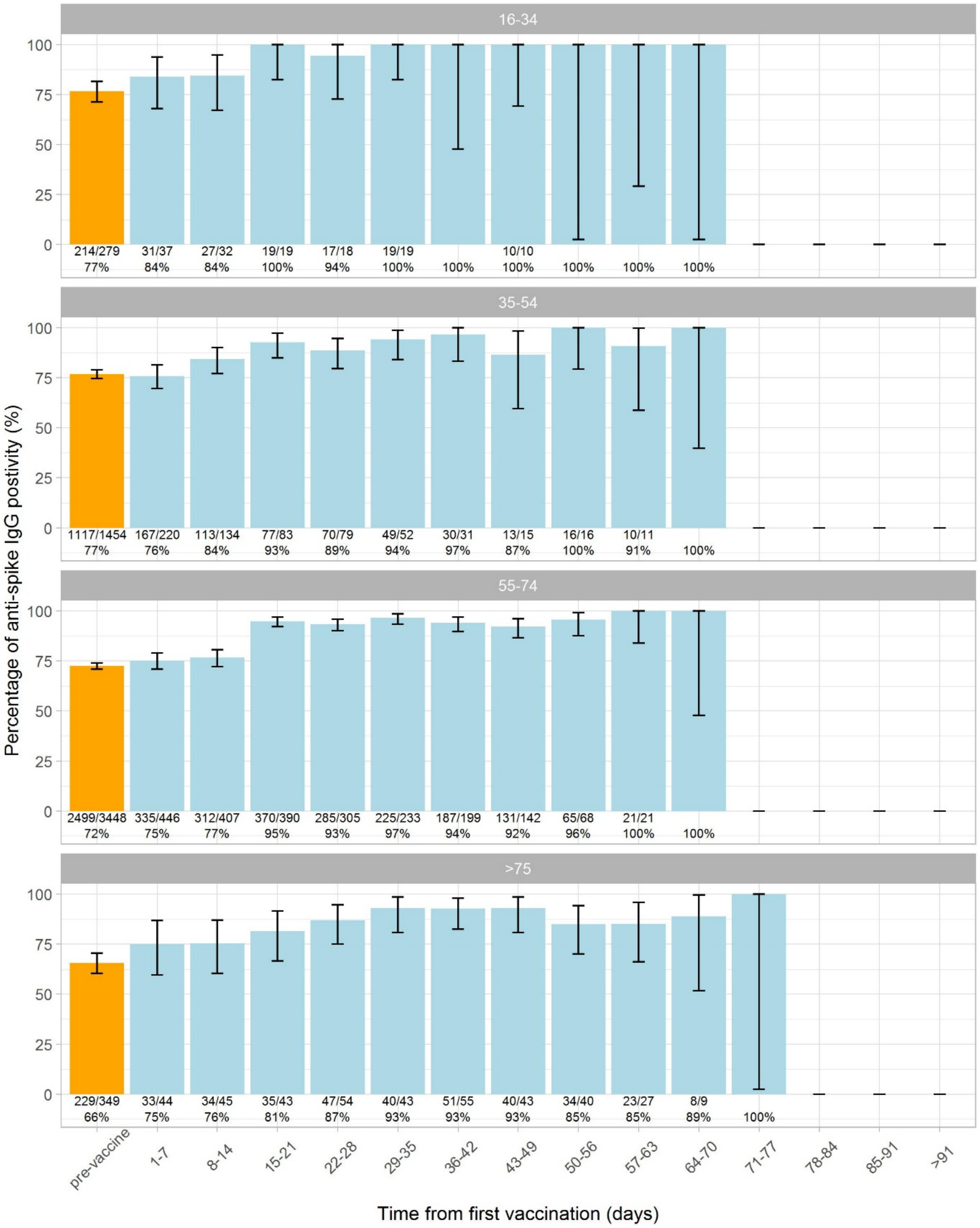


**Figure S4. Percentage (95% CI) anti-spike IgG positives by days after first vaccination in participants receiving a single dose of Oxford-AstraZeneca vaccine and with evidence of prior infection.** Results were divided into four age groups: 16-34, 35-54, 55-74, and >75 years.

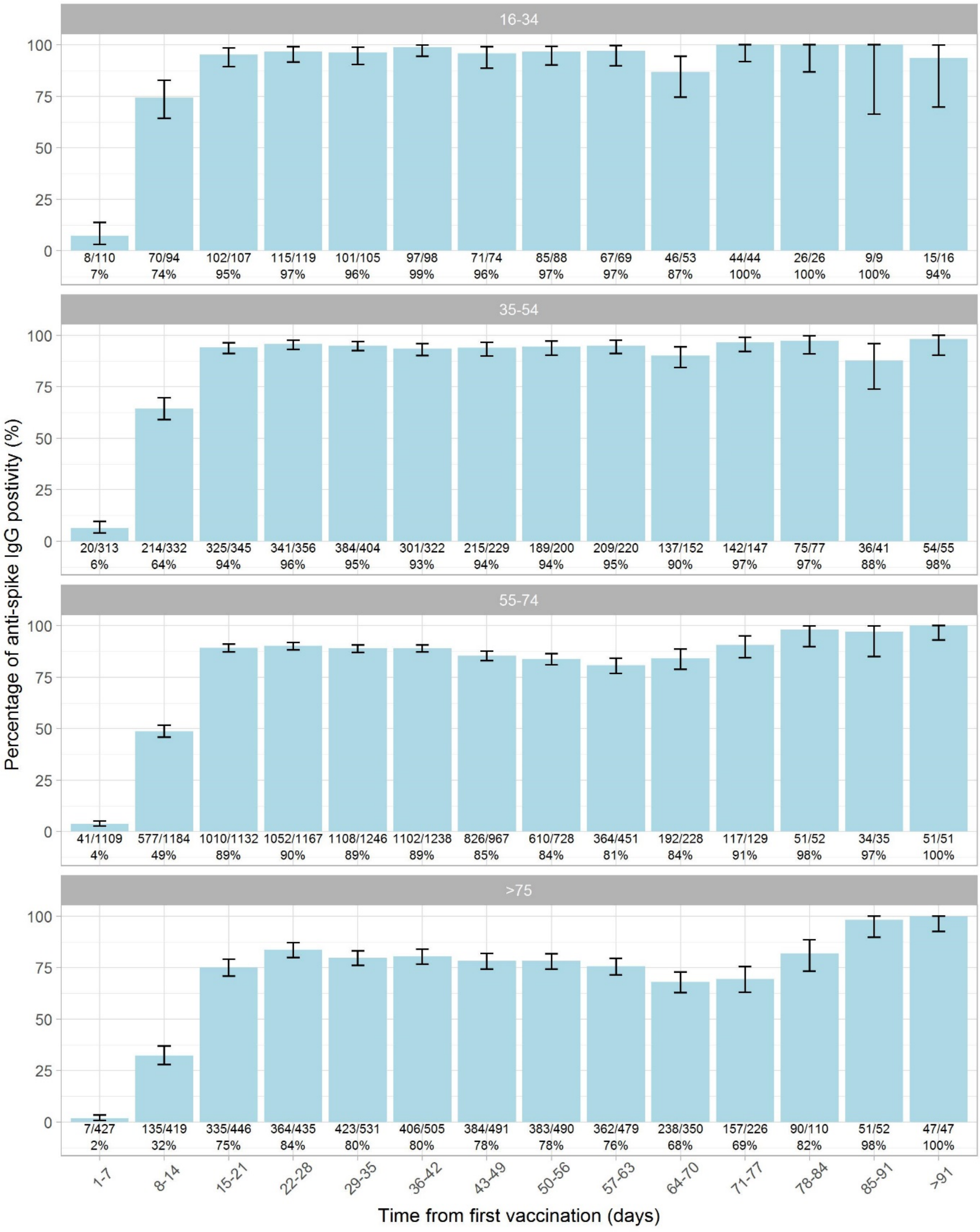


**Figure S5. Percentage (95% CI) anti-spike IgG positives by days after first vaccination in people receiving a single dose of Pfizer-BioNTech vaccine and without evidence of prior infection.** Results were divided into four age groups: 16-34, 35-54, 55-74, and >75 years.

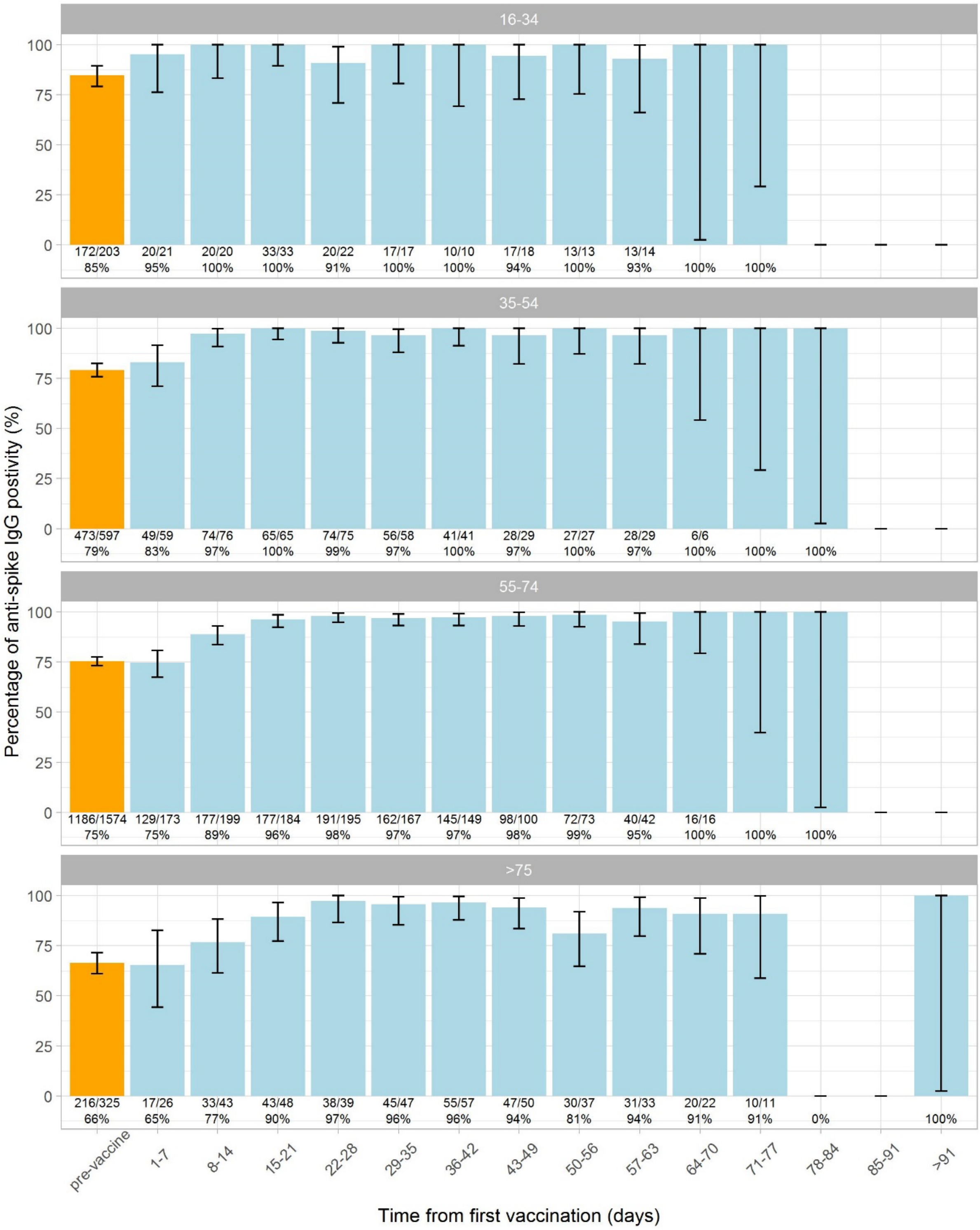


**Figure S6. Percentage (95% CI) anti-spike IgG positives by days after first vaccination in people receiving a single dose of Pfizer-BioNTech vaccine and with evidence of prior infection.** Results were divided into four age groups: 16-34, 35-54, 55-74, and >75 years.

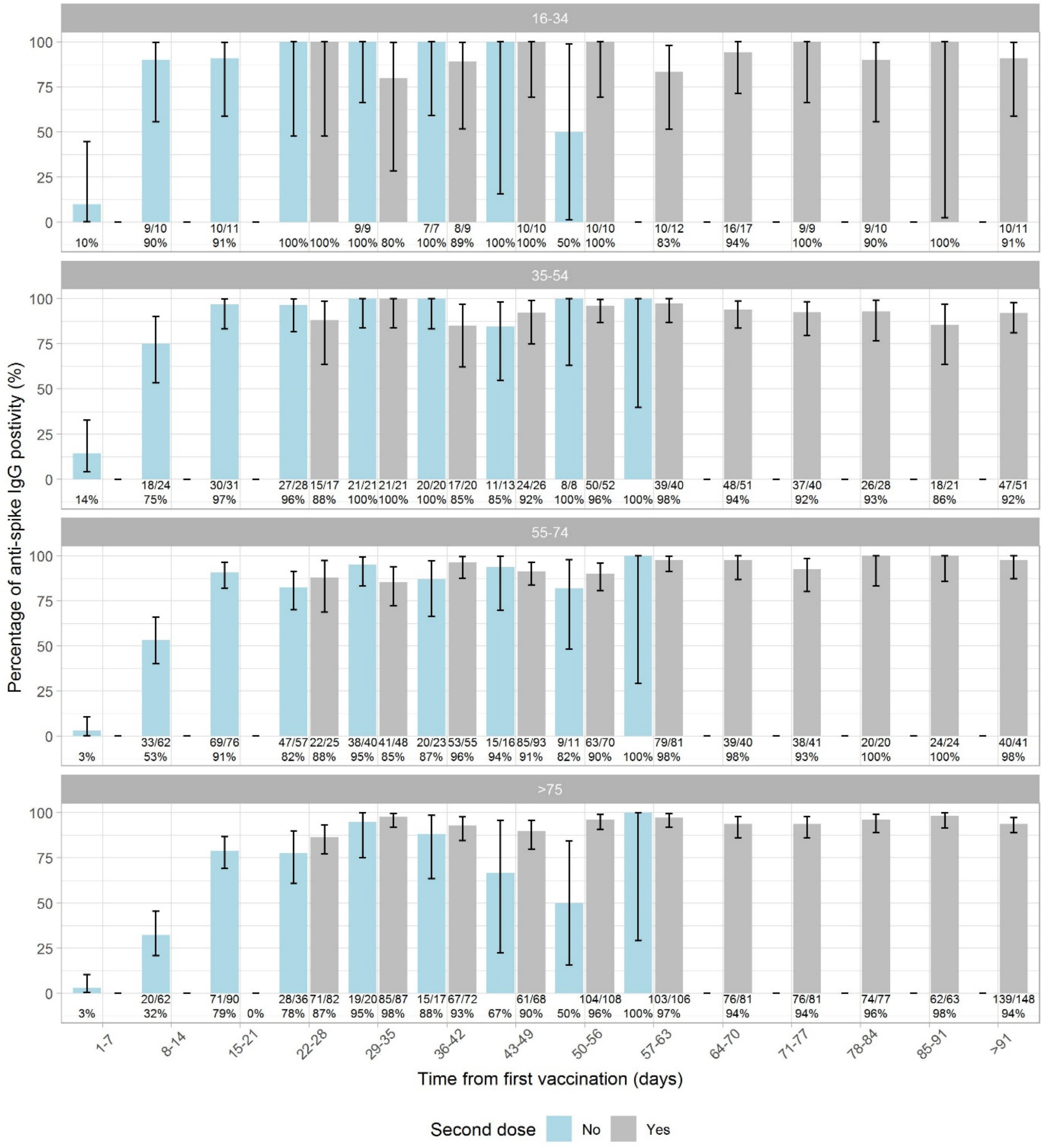


**Figure S7. Percentage (95% CI) anti-spike IgG positives by days after first vaccination in people receiving two doses of Pfizer-BioNTech vaccine and without evidence of prior infection.** Results were divided into four age groups: 16-34, 35-54, 55-74, and >75 years.

.

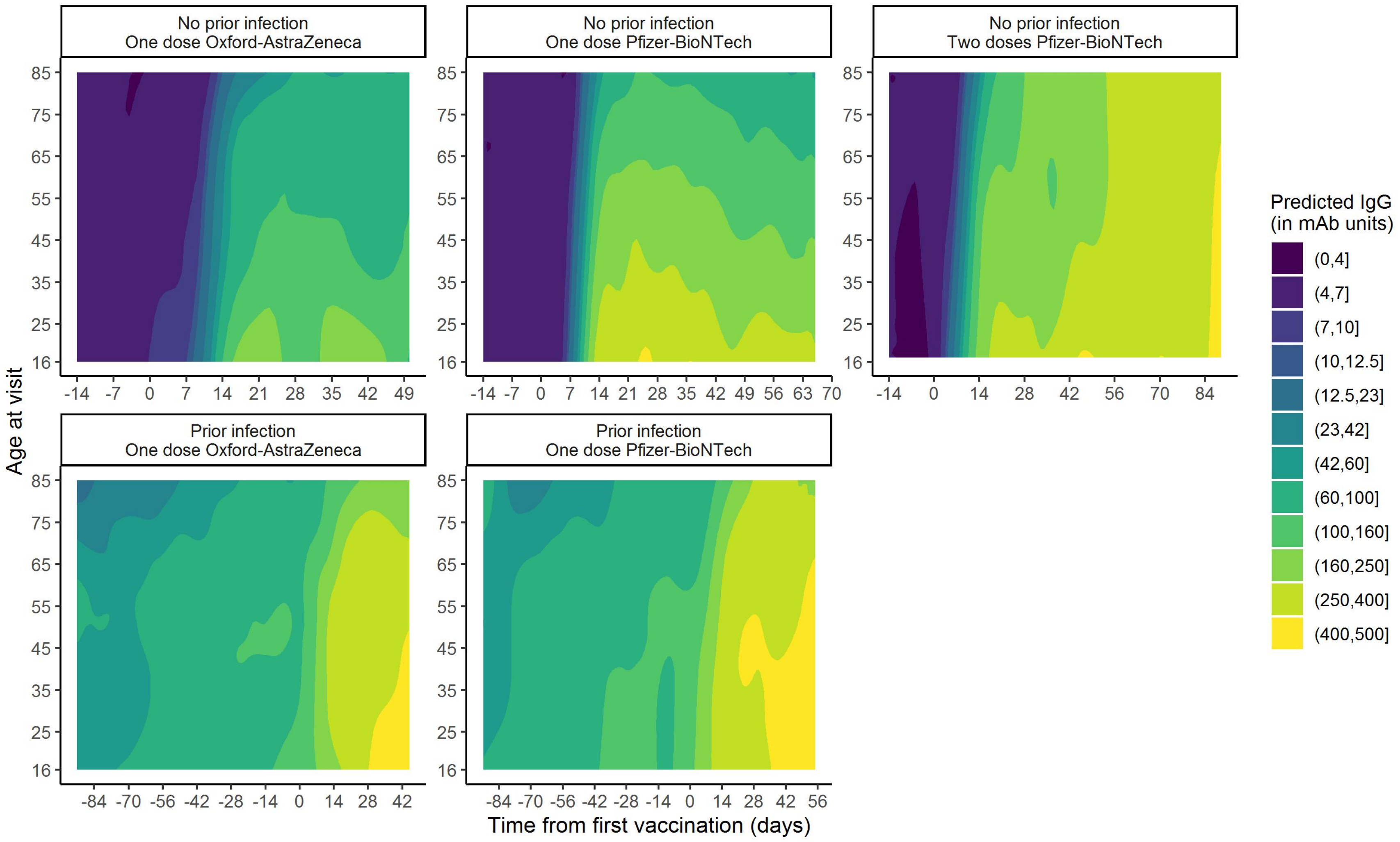


**Figure S8. Predicted anti-spike IgG levels (in mAb units) by time from first vaccination and age, according to vaccine type and prior infection status (full model).** Predictions shown for specific ages in **Figure 3.**

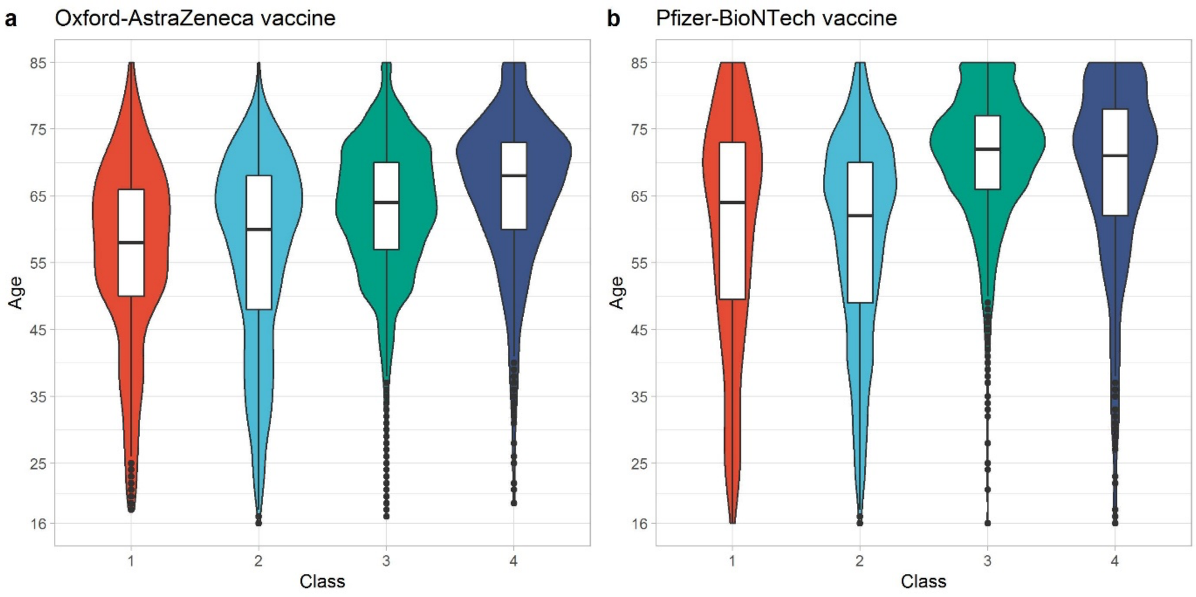


**Figure S9. Age distribution by classes identified from latent class mixed models for single dose Oxford-AstraZeneca and Pfizer-BioNTech vaccines.** 1='plausibly previously infected' group, 2='high response' group, 3='medium response' group, 4='low response group'. Same area for each violin, see Table S3 for numbers in each group.
